# T cell differentiation drives the negative selection of pathogenic mtDNA variants

**DOI:** 10.1101/2023.04.26.23289145

**Authors:** Imogen G. Franklin, Paul Milne, Jordan Childs, Róisín M. Boggan, Isabel Barrow, Conor Lawless, Gráinne S Gorman, Yi Shiau Ng, Matthew Collin, Oliver M. Russell, Sarah J. Pickett

**Author notes:** Corresponding authors: Sarah Pickett: Wellcome Centre for Mitochondrial Research, 4th Floor Cookson Building, Medical School, Framlington Place, Newcastle-upon-Tyne, NE2 4HH, UK.; E-mail., Matthew Collin: Haematopoiesis and Immunogenomics Laboratory, Room M3.127, 3^rd^ Floor Leech Building, Medical School, Framlington Place, Newcastle-upon-Tyne, NE2 4HH, UK. denote equal contributions (Imogen G. Franklin, Paul Milne, Matthew Collin, Oliver M. Russell, Sarah J. Pickett). Conflict of Interest: The authors have declared that no conflict of interest exists.

## Abstract

Pathogenic mitochondrial (mt)DNA single nucleotide variants are the most common cause of adult mitochondrial disease. Whilst levels of the most common heteroplasmic variant (m.3243A>G) remain stable in post-mitotic tissues, levels in mitotic tissues, such as blood, decrease with age. Given differing division rates, longevity and energetic requirements within haematopoietic lineages, we hypothesised that variant level decline is driven by cell-type specific mitochondrial metabolic requirements. To address this, we coupled cell sorting with mtDNA sequencing to investigate mtDNA variant levels within progenitor, myeloid and lymphoid lineages from 26 individuals harbouring pathogenic mtDNA variants. We report that whilst the level of m.3243A>G declines with age in all analysed cell types, the T-cell lineage shows a significantly greater decline. This was confirmed for a second pathogenic tRNA variant; m.8344A>G, indicating that this phenomenon is not limited to m.3243A>G. High-throughput single cell analysis revealed that decline is driven by increasing proportions of cells that have cleared the variant genome, following a hierarchy that follows the current orthodoxy of T-cell differentiation and maturation. This work identifies the unique ability of T-cell subtypes to selectively purify their mitochondrial genomes, and identifies pathogenic mtDNA variants as a new means to track blood cell differentiation status.

## Introduction

Mitochondrial diseases, characterised by impaired oxidative phosphorylation (OXPHOS), can be caused by pathogenic variants in either the mitochondrial or nuclear genomes. The most common cause of mitochondrial disease in adults is the mitochondrial DNA (mtDNA) m.3243A>G variant(1), which is located in the *MT-TL1* gene and primarily causes complex I (NADH: ubiquinone oxidoreductase) dysfunction via disruption of translation(2). Carriers present with a range of clinical phenotypes from asymptomatic to a severe neurological condition known as MELAS (Mitochondrial Encephalomyopathy, Lactic Acidosis, and Stroke-like episodes) syndrome. As mtDNA exists in multiple copies per cell, mixed wild-type and variant populations can co-exist, giving rise to heteroplasmy, which varies between individuals, tissues and cells; therefore, the relationship between tissue variant levels and both disease severity and phenotypic presentation is complex(3–7).

Unlike the majority of heteroplasmic mtDNA point mutations, there is strong evidence for negative selection against m.3243A>G mtDNA molecules within mitotic tissues, particularly in blood(3, 4, 6, 8–11). Blood represents an ideal tissue to study selection pressure as it is composed of multiple different cell types within the same microenvironment and is easily accessible. Previous modelling results are consistent with a decline initiating in haemopoietic stem cells, with a mechanism involving the random segregation of mtDNA molecules at mitosis followed by the elimination of cells that have reached a biochemical threshold due to the accumulation of high levels of variant mtDNA (10). More recently, an enhanced decline of m.3243A>G within T cells has been demonstrated(12); this is consistent with a proposed mtDNA bottleneck within lymphoid cell development(13). However, this study did not analyse progenitor cells, and so the question of how progress through the lymphoid differentiation bottleneck drives m.3243A>G decline remains.

Given differing division rates, longevity and energetic requirements within both the myeloid and lymphoid compartments, we hypothesised that mutation level would be determined by development stage within each haematopoietic lineage. To address this, we coupled fluorescently activated cell sorting (FACS), which allowed a finer resolution of cellular phenotype, with a novel, high-throughput single cell mtDNA sequencing technique to investigate m.3243A>G mutation load within thirteen blood cell compartments. We compared this with a second variant, m.8344A>G, which to-date has not been reported to decline with age(14). We show that mtDNA mutation load tracks with T cell differentiation status, with less mature, naïve cells exhibiting higher mutations loads.

## Results

### 1. Mitochondrial disease patients have a lower proportion of T cells

Blood samples from 22 patients genetically confirmed to harbour m.3243A>G and 16 controls were fractionated by density centrifugation followed by FACS to separate whole blood into a broad range of cell types of differing maturities (n=13; **Figure 1, S1**). We first investigated whether the presence of m.3243A>G affected cellular proportions. We found that individuals carrying m.3243A>G have significantly lower proportions of T cells (p=0.0001) and a higher proportion of CD3-HLA-DR+ antigen presenting cells and B cells (p=0.0001 and p=0.0045, respectively) than controls (**Figure 2a**). Within the T cell compartment, patients and controls have similar CD4+:CD8+ and naïve:memory ratios (**Figure 2bi**). There were also no differences in naïve:memory ratios for B cells and CD56 bright/dim for NK cells (**Figure 2bii**). Monocyte and dendritic cell proportions do not differ between patients and controls (**Figure 2c**).

**Figure 1:**
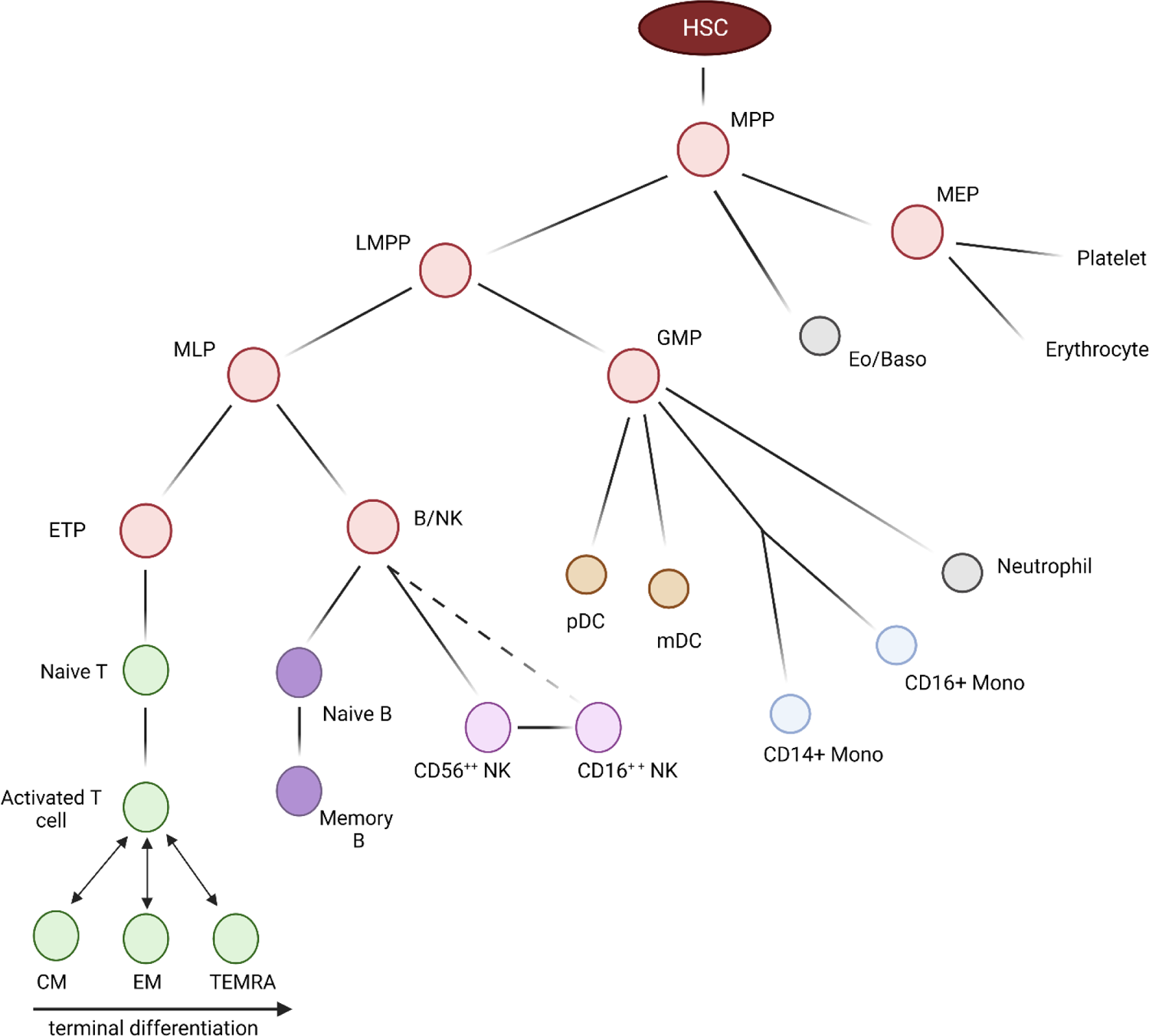
Investigated blood cell types. Haematopoietic differentiation: Blood cells are derived from haematopoietic stem cells in the bone marrow; these give rise to highly proliferative CD34+ progenitors (red) which occur infrequently in peripheral blood. Monocyte (blue), granulocytes (grey), dendritic (orange) and Natural Killer (NK; light purple) cells are involved in the innate immune response. B (dark purple) and T (green) lymphocytes communicate with these cells to generate the highly specific adaptive immune response; B and T memory cells remain after clearance of a pathogen to defend against reinfection. Dashed line represents differentiation yet to be validated. HSC, haematopoietic stem cell; MPP, multipotent progenitors; LMPP, lymphoid-primed multipotent progenitors; MLP, multilymphoid progenitors; ETP, early T-cell precursor; B/NK, B & NK cell progenitor GMP, granulocyte-macrophage progenitors; MEP, megakaryocyte-erythroid progenitor; Eo/Baso, eosinophil/basophil; CM, central memory T-cell; EM, effector memory T-cell; TEMRA, terminal effector cell (CD45RA+ effector memory T-cell); NK, natural killer cell; pDC, plasmacytoid dendritic cell; mDC, myeloid dendritic cell. Created with BioRender.com

**Figure 2:**
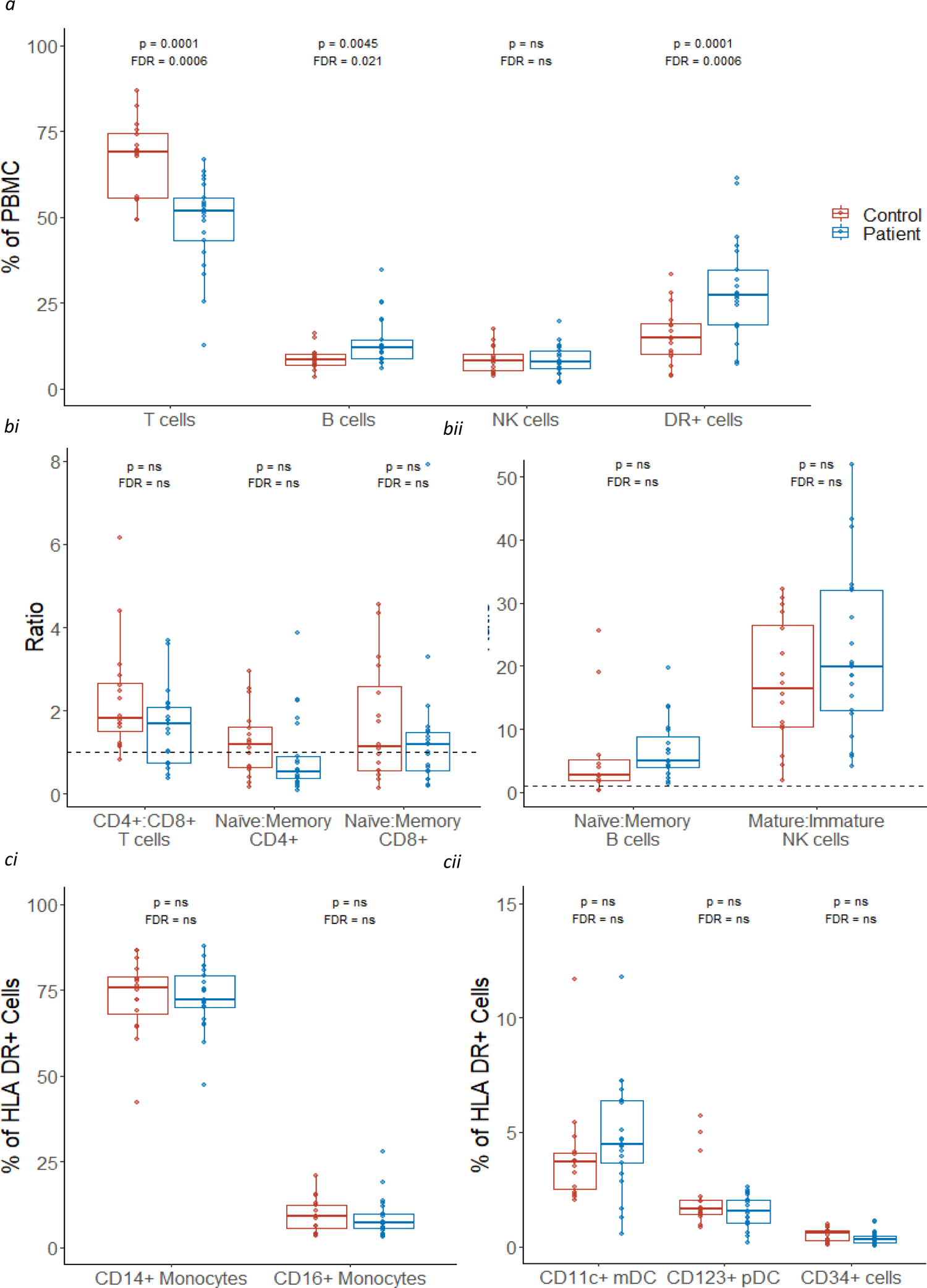
Comparison of the proportions of cellular subsets in patients and controls. Percentage quantification for each cellular subset was derived from flow cytometry data for 21 patients (blue) and 16 controls (red); data were unavailable for P12. See materials and methods and for gating strategy. Values in patients and controls were compared using linear regression, controlling for age at sample; both p values (upper) and values adjusted for false discovery rate (FDR; lower) for all comparisons in the figure are shown; ‘ns’ denotes values >0.05. a) Proportion of T, B, NK and HLA DR+ cells. b) Ratio of CD4+:CD8+ T cells, and naïve:memory CD4+ and CD8+ T cells, B cells and NK cells; a ratio of 1 is represented with a dotted line. c) Proportion of subsets of HLA CD+ cells. DR+ cells: HLA DR+ cells CD34+: Precursor cells NK: natural killer cells; CD11c+ mDCs: CD11c+ myeloid dendritic cells; CD123+ pDCs: CD123+ plasmacytoid dendritic cells; PBMC: peripheral blood mononuclear cells.

### 2. Selection against m.3243A>G is strongest within the memory T cell compartment

To understand the cell-type-specific relationship of m.3243A>G levels with age, we measured m.3243A>G levels within these blood cell populations from 22 patients. All cellular populations studied showed a significant age-associated decline in m.3243A>G levels (p value range = <0.0001 - 0.0012; **Figure 3a**).

**Figure 3:**
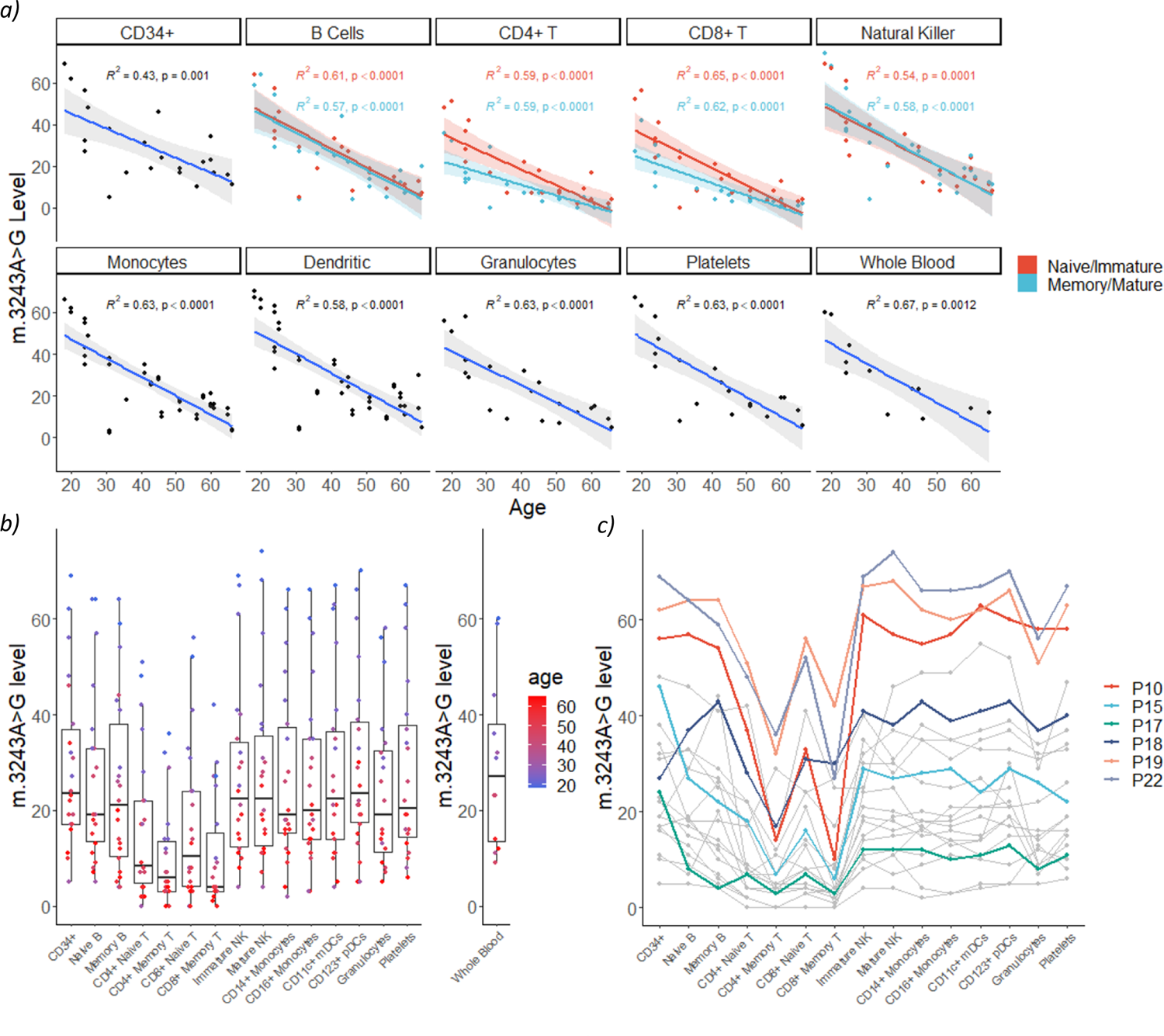
m.3243A>G levels across bulk populations of peripheral immune cells. Thirteen blood cell populations were separated by flow cytometry (n = 22); levels in platelets (n=20) and granulocytes (n=20) separated using density centrifugation from the same whole blood sample are also shown. Each point in the flow cytometry-derived cell populations represents m.3243A>G levels in ∼1000 cells isolated from a single individual. a) m.3243A>G level is negatively correlated with age in all cell populations studied; each point represents m.3243A>G level in a cell population from one individual. Naïve / immature cell populations are represented in blue and memory /mature populations in red. Lines represent linear models with 95% confidence intervals; adjusted R^2^ and p values are shown. Linear mixed models of the effect of age and maturity on m.3243A>G levels including a term for the interaction of age with maturity show that m.3243A>G levels are lower in memory vs. naïve CD4+ and CD8+ T cells (p=0.0034, p=0.0011) but not B cells (p=0.7370) or mature vs. immature NK cells (p=0.5275). The slope is also significantly less steep for naïve CD4+ and CD8+ T cells (p=0.0411 and p=0.0223) but not for B or NK cells (p=0.9898 and p=0.6146). b) All cell types studied within the T cell compartment have significantly reduced m.3243>G levels compared to CD34+ precursor cells, the most naïve blood cells investigated (linear mixed model accounting for patient as a fixed effect; CD4+ Memory: β=-19.55, SE=1.66, p<0.0001; CD8+ Memory: β=-18.82, SE=1.666, p<0.0001; CD4+ Naïve: β= 12.82, SE=1.66, p<0.0001; CD8+ Naïve: β=-12.18, SE=1.66, p<0.0001) as well as the memory B cells (β=-4.36, SE=1.66, p=0.0090) and granulocytes (β= −5.72, SE= 1.70, p=0.0009). Where available, m.3243A>G levels in whole blood are also shown (n=12) c) The same data represented with lines connecting points from individual patients. Colours represent samples from the six patients taken forward for single cell analysis. CD34+: Precursor cells; Immature NK: immature natural killer cells; mature NK: mature natural killer cells; CD11c+ mDCs: CD11c+ myeloid dendritic cells; CD123+ pDCs: CD123+ plasmacytoid dendritic cells.

Comparison of m.3243A>G levels in all cells relative to CD34+ haematopoietic progenitors, revealed an enhanced clearance of the m.3243A>G variant allele in all T cell subsets (reduction range=12.18-19.55%, p values<0.0001). This is consistent between both CD4+ and CD8+ T cells. Interestingly, naïve T cells had significantly higher m.3243A>G levels than their memory cell counterparts (**Figure 3a**; CD4+: p=0.0034; CD8+: p=0.0011); indicating that mutation levels further reduce as T cells proliferate and differentiate into memory cells. We didn’t see this maturity-associated enhanced decline in B or NK cell lineages (p values>0.05).

For both CD4+ and CD8+ T cells, negative correlations between age and mutation levels are higher in naïve cells compared to their memory counterparts (**Figure 3a**; interaction p=0.0411 and p=0.0223), consistent with weaker negative section at lower m.3243A>G levels. Interestingly, for one patient there was no detectable m.3243A>G present in any of the T cell populations studied (**Figure 3c**; P05, whole blood m.3243A>G level 6%) implying that although negative selection may be weaker, it is still present at low m.3243A>G levels.

We also detected a small reduction in mutation levels in memory B cells and granulocytes (**Figure 3b**; reduction range=4.36-5.72%; p=0.0090 and p=0.0009, respectively) compared to CD34+ progenitor cells. For the granulocytes, which were isolated by density centrifugation rather than FACS, we cannot rule out a small degree of contamination with other cell types including T cells which are the most abundant mononuclear cell. Unlike T cells, there was no significant difference between the levels in naïve and memory B cells.

### 3. mtDNA copy number decreases with age in mitochondrial disease patients

m.3243A>G is functionally recessive; low levels of wild-type mtDNA are capable of rescuing defective oxidative phosphorylation *in vitro*(15, 16) and higher mtDNA copy number (CN) in skeletal muscle is associated with a lower disease burden(17). Therefore, we sought to investigate the relationship between mtDNA CN and m.3243A>G in blood cells.

For most cell types studied, mtDNA CN is similar between m.3243A>G patients and controls (**Figure 4a**; Wilcoxon tests; p>0.05), with the exception of CD11c+ myeloid DCs (p=0.0186), although this was not significant after correcting for multiple testing (FDR-adjusted = 0.2418). Interestingly, mtDNA CN is negatively correlated with age in cells from m.3243A>G patients (**Figure 4b**; linear mixed model; β = −0.0121, SE = 0.0031, p = 0.0017) but there is no relationship in controls (p=0.2133). Given the relationship between age and m.3243A>G level, this is likely to reflect a positive correlation between m.3243A>G level and mtDNA CN (**Figure 4c**), suggesting that cells respond to higher m.3243A>G levels by increasing mitochondrial biogenesis. Analysis stratified by cell type reveals that this is likely to be driven by cells of lymphoid origin; mtDNA CN is negatively associated with age and positively associated with m.3243A>G level in patient cells of lymphoid origin (**Figure S2**; CD4+ T cells, CD8+ T cells, B cells and NK cells). In T cells, the relationship with mutation level is higher (beta ranges; T cells: 0.0229-0.0234; B and NK cells: 0.0096-0.0106), indicating that T cells are more sensitive to the negative effects of m.3243A>G than other blood cell types.

**Figure 4:**
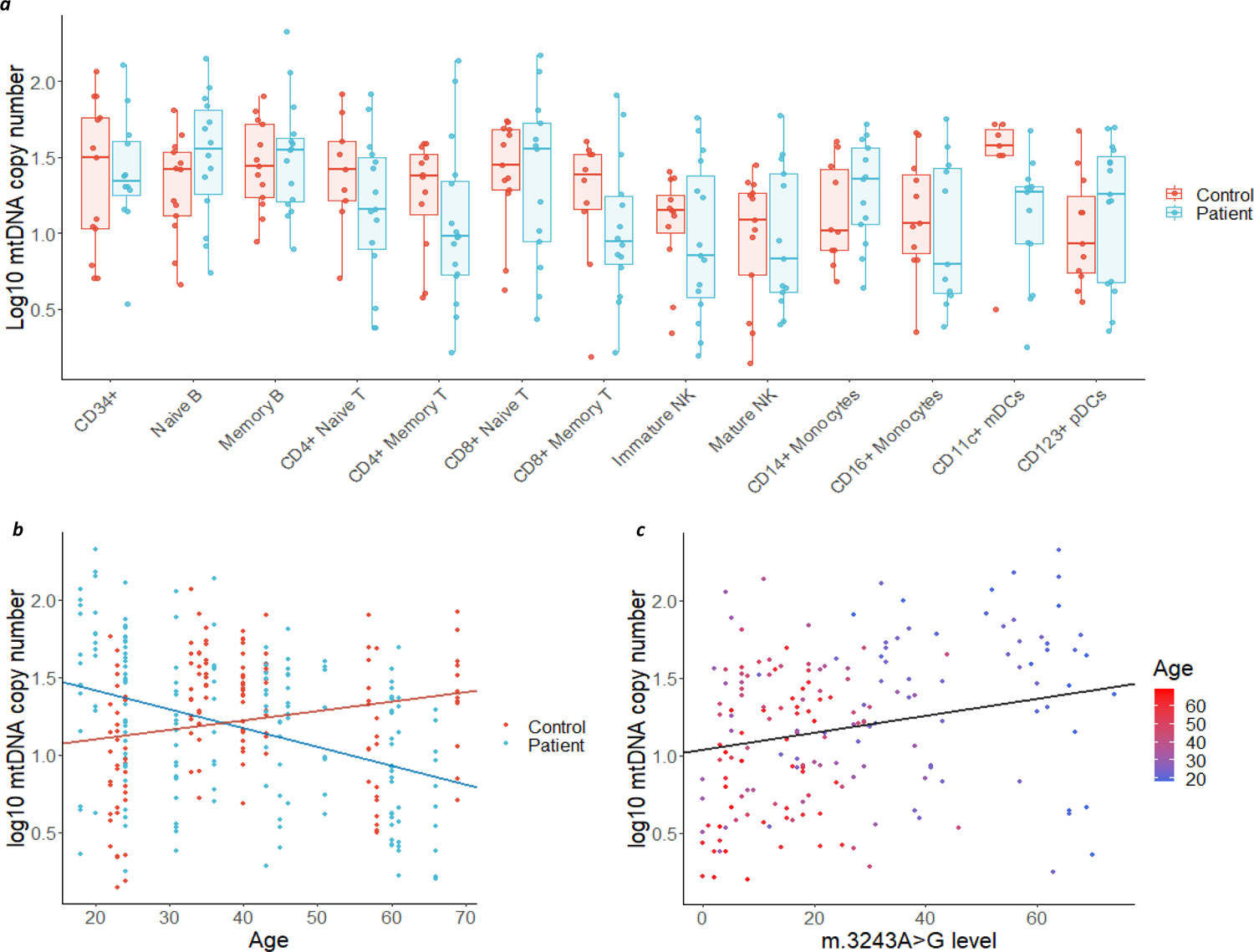
mtDNA copy number in bulk populations of peripheral immune cells. Each point represents relative per cell mtDNA copy number derived from ∼1000 cells from a population of cells, separated using flow cytometry, from a single individual. a) Comparison of log mtDNA copy number between patients (blue; n = 16) and controls (red; n = 14). mtDNA CN is higher in controls than patients in CD11c+ mDCs (Wilcoxon test; p=0.0186, FDR-adjusted = 0.2418) but not in other subsets (p>0.05). b) Age and log mtDNA copy number are negatively correlated in m.3243A>G patients (linear mixed model; β = −0.0121, SE = 0.0031, p = 0.0017) but not in controls (β = 0.0060, SE = 0.0046, p = 0.2133). c) m.3243A>G level and log mtDNA copy number are weakly positively correlated (linear mixed model; β = 0.0054, SE = 0.0024, p = 0.0263). Points are coloured by age; as m.3243A>G level in the blood declines with age, the effect of age is difficult to distinguish from m.3243A>G level.

### 4. Selection against pathogenic mtDNA variants in the T cell compartment is not limited to m.3243A>G

To understand whether this enhanced selection within the T cell compartment is specific to m.3243A>G, we carried out similar investigations in four individuals carrying the m.8344A>G variant, which is located within *MT-TK* (encoding tRNA^lys^) and has been previously reported to maintain stable levels in the blood with age(14, 18, 19).

Consistent with our findings from m.3243A>G, levels of m.8344A>G are significantly reduced in memory T cells (CD4+ and CD8+; p<0.0001), memory B cells (p=0.0038) and granulocytes (p=0.0400) when compared to CD34+ progenitors (**Figure 5**). Although we observe the same trend within naïve T cells, this is not significant.

**Figure 5:**
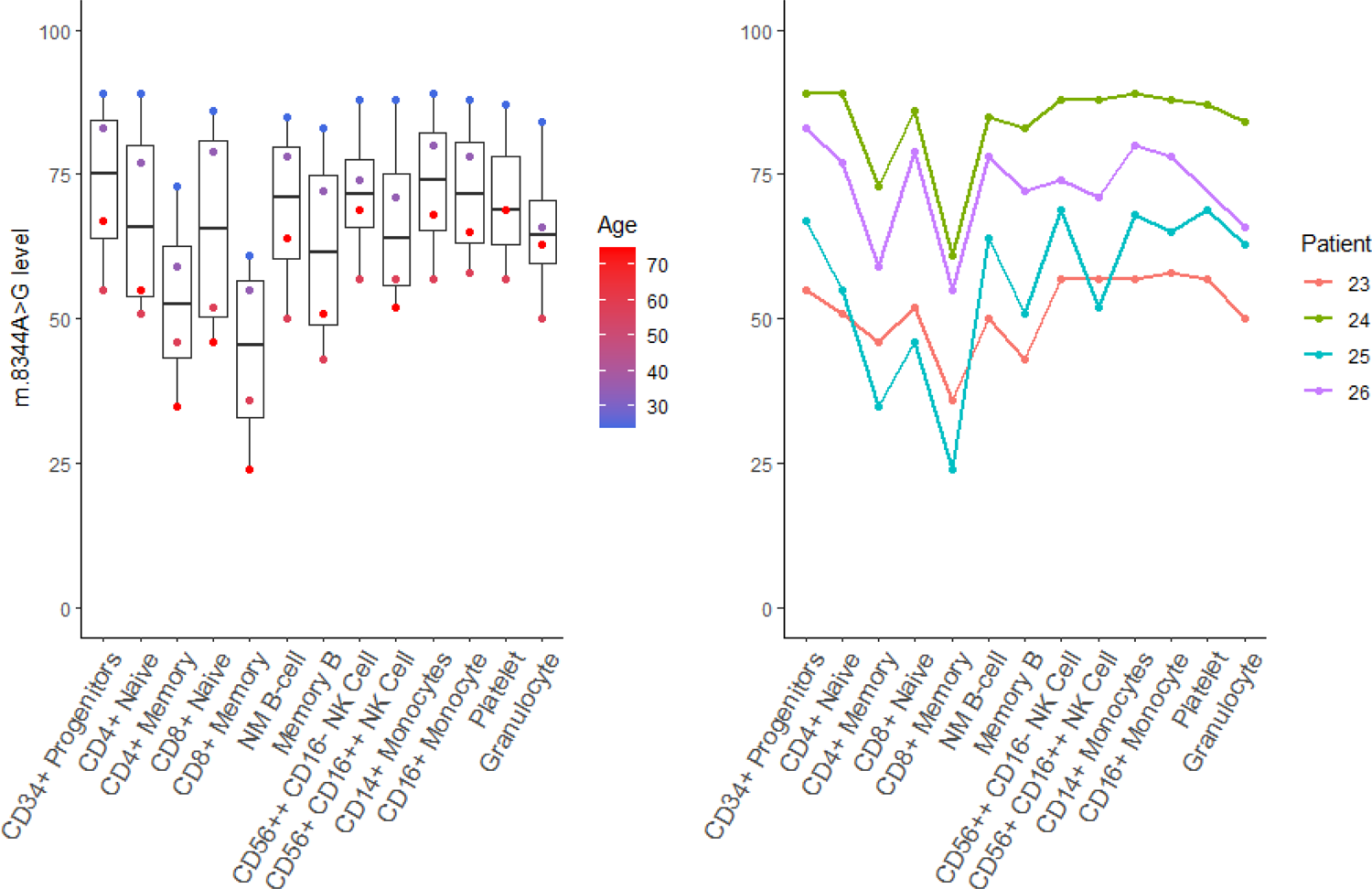
m.8344A>G levels across bulk populations of peripheral immune cells. Eleven blood cell populations were separated by flow cytometry (n = 4); levels in platelets and granulocytes (n=4) separated using density centrifugation from the same whole blood sample are also shown. Each point in the flow cytometry-derived cell populations represents m.3243A>G levels in ∼1000 cells isolated from a single individual as detailed in figure 1. Memory lymphoid cells as well as granulocytes possess a m.8344A>G-level significantly reduced from the CD34+ precursor cells (linear mixed model accounting for patient as a fixed effect; CD4 memory: β=-20.25, SE=3.63, p<0.0001; CD8 memory T: β=-29.5, SE=3.63, p<0.0001; memory B cells: β=-11.25, SE=3.63, p=0.0038; granulocytes: β=-7.75, SE=3.63, p=0.0400).

### 5. Single cell analysis reveals cells that have reverted to wild-type

Having identified an enhanced reduction in m.3243A>G levels within T cell populations, and a further reduction particularly within the memory T compartment, we sought to understand the dynamics of this by studying m.3243A>G levels within single cells from six patients (age range = 18-46, whole blood m.3243A>G level range = 9-60%). To achieve this, we devised a novel, high throughput, targeted next generation sequencing assay, allowing us to determine m.3243A>G level within 5732 single cells which had been sorted by FACS into each specific cell population (**Figure S3)**. To gain insight into this enhanced negative selection within the T cell compartment, we further subdivided memory T cells into effector memory (EM), central memory (CM) and TEMRA cells (effector memory T cells re-expressing CD45RA) (gating strategy, **Figure S1**).

For most cell populations investigated, the range of single cell m.3243A>G levels spans almost the entire possible range (**Figure 6**; range=0.00-99.23%). This is more obvious in cells from younger patients but is also evident in the majority of cell types for P15 (age range = 36-45 years). Despite a much lower whole blood mutation level for P17 (12% at 41 years), four of the nine cell subsets for P17 (age range = 46-55 years) have cells with >65% m.3243A>G. These single cell data show that the enhanced negative selection against m.3243A>G in T cells can be explained by an increased number of cells with near zero levels of mutation, with a trend towards total clearance, rather than a general reduction in levels across all cells. This implies that selection against the G allele continues to occur even at very low mutation levels.

**Figure 6:**
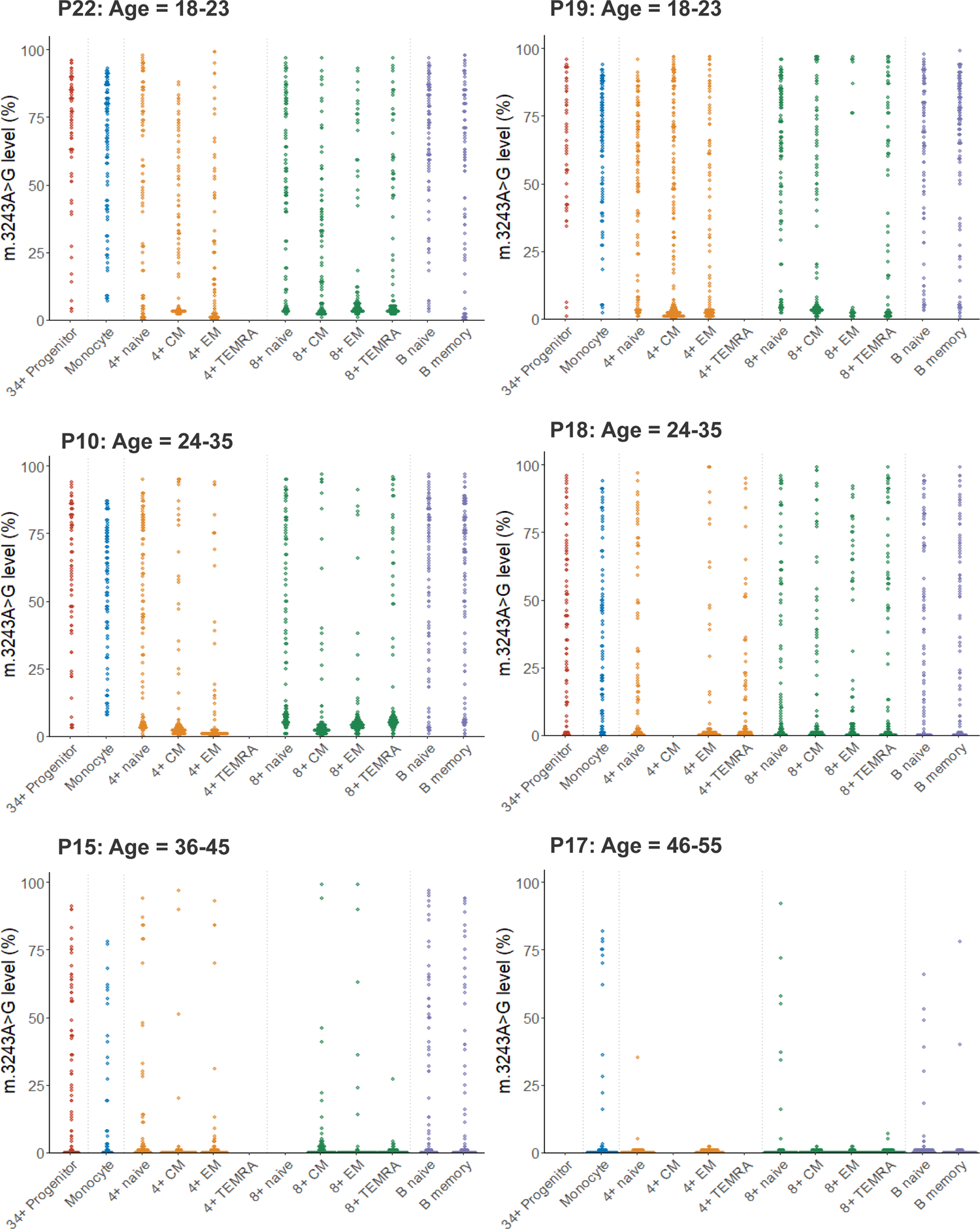
Dot plots showing single cell distributions of m.3243A>G levels in peripheral immune cell subsets for six patients. Each point represents the m.3243A>G level in a single cell from a single cell subtype from a single individual (median number of single cells in each subset=91; IQR=87, 93, range=26, 200). Colours represent broad cell subtypes (red: CD34+ precursor cells; blue: monocytes; orange: CD4+ T cells; green: CD8+ T cells; purple: B cells). Where possible, T cells were further separated into naïve, effector memory (EM), central memory (CM) and TEMRA populations and B cells into naïve and memory cells (see Supplementary figure XX for gating strategy). Sufficient CD4+ TEMRA cells were only present in one patient (P18). Data for some cellular subsets were removed during data quality control and are shown as missing data (P18: CD4+ CM; P15: CD8+ naïve; P17: CD34+ precursors, CD4+ CM; see Materials and Methods for quality control details).

### 6. Enhanced negative selection in all subsets of memory T cells

To determine the proportion of cells that have cleared m.3243A>G within each cell population, we quantified the proportion of cells with a near zero level of m.3243A>G using Bayesian inference (**Figure 7, Table S6**). CD34+ progenitors and monocytes had similar proportions of cells that had cleared m.3243A>G and this proportion increased with age; a pattern that was seen for all lineages (**Table S6**). Within the T cell compartment, the proportion increases with maturation; for both CD4+ and CD8+ cells, all memory cells had a significantly higher proportion of near-zero cells than the naïve cells in the majority of individuals. Therefore, selection against m.3243A>G continues during the transition from naïve to memory T cell.

**Figure 7:**
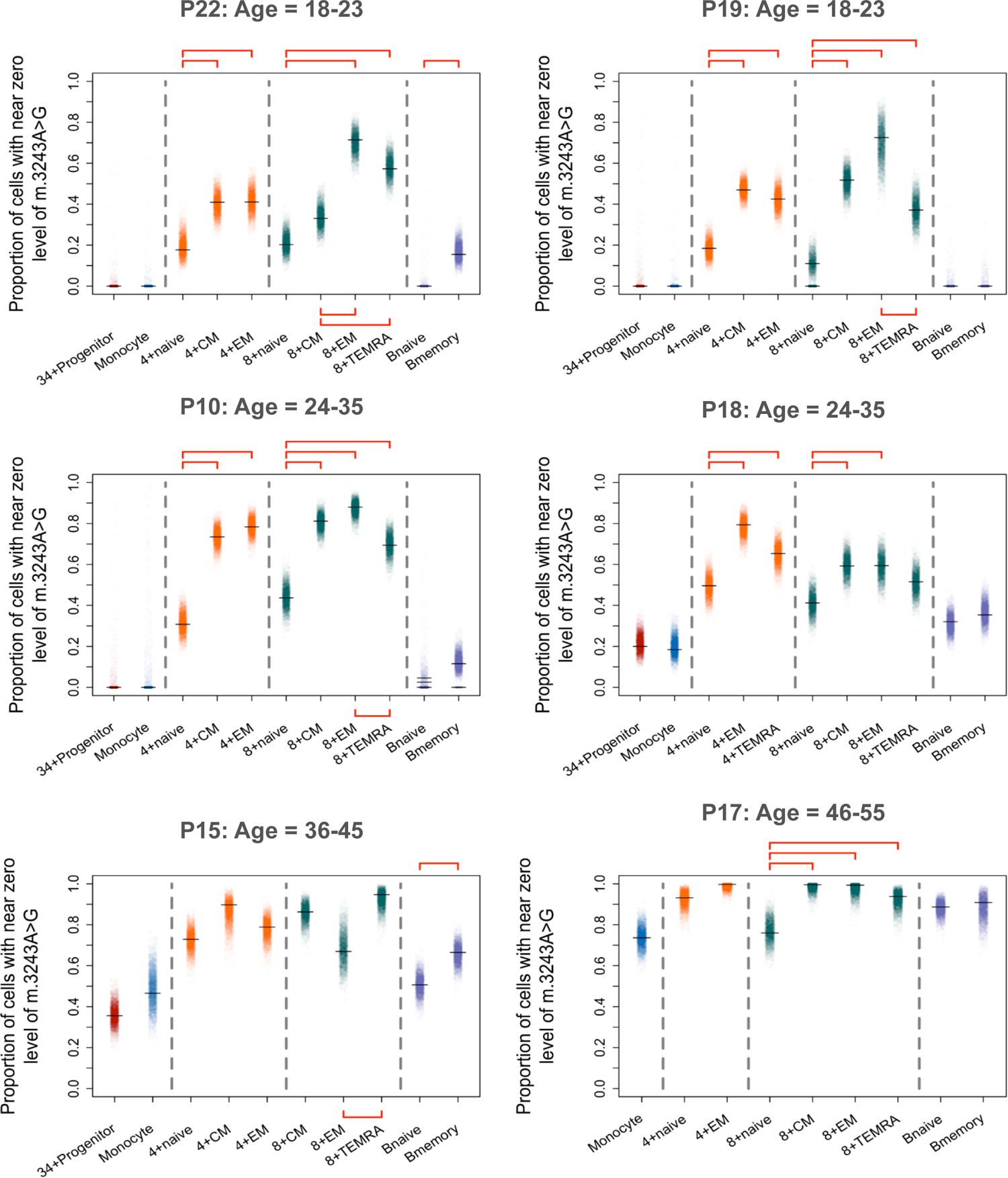
Proportion of cells that have cleared m.3243A>G increases with cell maturity. Each point represents the estimate from one Bayesian iteration. Peak estimates are indicated with horizontal black lines; multiple lines are shown when the posterior distribution is bi/multi-modal. Significance testing was carried out between cell-type pairs within major cell-type groups (indicated with horizontal dotted lines); red horizontal bars indicate pairs of estimates that are considered statistically different (i.e. zero lies outside the 95% credible interval of the posterior difference between the proportions; see methods for details).

Results comparing CM and EM cells were varied; CD4+ EM and CM cells showed a similar proportion of near zero cells, whereas CD8+ EM and CM cells were more variable. CD8+ EM had a larger proportion than CD8+ CMs in 2/6 individuals (one of which was significantly different), but in 3/6 individuals the proportions were similar or showed the opposite trend (**Figure 7, Table S6**).

Interestingly, in 5/6 individuals, CD8+ TEMRA cells had a lower proportion of cells that had cleared the variant allele than EMs; this difference was significant in 3/6 individuals. B memory cells also have a higher proportion of cells that have cleared the variant allele than naïve B cells; although this difference with maturity is not as clear or as large as that seen in T cells, the trend is present across all individuals and is significant in 2/6 individuals. Notably, the most mature B cell, the plasma cell, is not found in peripheral blood.

## Discussion

Selection against the m.3243A>G variant in the blood tissue is widely recognised in the literature but the mechanism by which this occurs is unknown(3, 6, 8–11). Our analysis reveals that this phenomenon is not limited to m.3243A>G, occurs in cells of both lymphoid and myeloid origin, as well as circulating CD34+ progenitor cells (the earliest cell in haematopoietic differentiation that was investigated), and is particularly enhanced within the T cell compartment. These results are consistent with research by Walker and colleagues who, via ATAC-seq, demonstrated a significantly increased proportion of lymphocytes with low m.3243A>G levels(12). By using FACs, to achieve a finer resolution of differentiation trajectories, coupled with a customised high-throughput single cell mtDNA sequencing assay, we show that this phenomenon is enhanced within central and effector memory T cells, as well as TEMRA cells, demonstrating that the proportion of cells with negligible heteroplasmy continues to increase throughout the T cell lifespan. The trend we observe in the proportion of cells that cleared the mutation supports the conventional view of T cell differentiation, although we were surprised to find that TEMRA cells had a lower proportion of cells that had cleared the variant than EMs, implying either that TEMRA cells are less sensitive to m.3243A>G or that they are not directly derived from EM cells, according to alternative models of T cell development(20).

Our data also show, for the first time, a reduction in the relative abundance of T cells in m.3243A>G carriers compared to controls, suggesting the presence of this variant specifically impacts T cell homeostasis, the lineage in which the greatest selective pressure was observed. The difference in T-cell abundance remained significant after removal of data from P1, the only patient who had an abnormal lymphocyte count **(Table S3)**. The higher proportion of CD3-HLA-DR+ cells we observed could simply reflect the lower proportion of T cells, as these are the two largest compartments within PBMC. The link between mitochondrial disease and the immune system is not fully understood but recurrent infections have been reported in some patients with mitochondrial disease and can result in a severe inflammatory response(21). Interestingly, we did not see a difference in the proportion of T-cells in m.8344A>G patients compared to controls; a larger m.8344A>G cohort is needed to elucidate this further.

T cell development involves a rigorous central tolerance process within the thymus, which involves rapid cell proliferation followed by death of unreactive/autoreactive cells(22). A further proliferative expansion occurs within circulating T cells in response to infection, after which, antigen-experienced memory T cells are poised to mount a response to reinfection(23). Proliferative episodes followed by apoptosis or senescence of cells that have developed a respiratory complex deficiency due to the accumulation of a high mutation load are likely to impact m.3243A>G level distribution within a population(10, 24–26). However, respiratory complex deficiency can be rescued with as little as 6% wild-type mtDNA(16), and estimates for the threshold m.3243A>G level in muscle fibres are >80%(27). It is therefore puzzling that T cells clear the pathogenic variant to levels below the accepted threshold for a biochemical defect for post-mitotic tissues. Thus, the mechanism underlying the decline of m.3243A>G, and perhaps other pathogenic mtDNA variants, is either the result of unique metabolic constraints endured by immune cells or not wholly attributable to OXPHOS deficiency at a single cell level. Statistical modelling of m.3243A>G decline is consistent with selection occurring at the stem cell level(10); our data demonstrate that levels in CD34+ progenitor cells are negatively correlated with age and so support this theory. The discovery that m.8344A>G shows an enhanced decline in selected lineages, despite relatively stable levels in whole blood(14), indicates that specific immune cells are more sensitive to m.8344A>G, or pass through more rounds of purifying selection than stem cells. This raises the question of whether T cell sensitivity is a general phenomenon amongst pathogenic mt-tRNA variants and could be related to defects in mitochondrial translation(28). Future work should expand this study to include a wide range of variants within mtDNA protein-coding genes as well as other mt-tRNA genes.

The relationship between a T cell and its mitochondria is complex; cellular metabolism within the T cell is modified throughout its lifespan and plays a key role in determining cell fate(29, 30). Both proliferating immature thymocytes and activated T cells rely heavily on glucose to regenerate ATP via glycolysis to support rapid proliferation, which might imply that selection against OXPHOS-deficient cells is low at this point. In contrast, circulating naïve and memory T cells, which turnover slowly, are more reliant on OXPHOS(23, 31). Only a subset of activated T cells differentiate into long-lived memory cells; their long-term cellular survival and function is uniquely dependent upon spare respiratory capacity, facilitated by enhanced fatty acid oxidation and associated with higher expression of complex I(32). Therefore, naïve T cells containing mtDNA mutations are likely to be compromised, perhaps setting the stage for becoming vulnerable to m.3243A>G induced mitochondrial dysfunction upon activation.

T cell activation is associated with an increased membrane potential(33). Inhibition of OXPHOS complexes I and IV, both of which show deficiency in the presence of m.3243A>G(2, 34–38), induce activation defects in both CD8+ and CD4+ T cells(33, 39–41). Additionally, inhibiting glycolysis shifts T cell fate away from senescence and towards a self-renewal phenotype(42). Therefore, glycolysis alone is insufficient to sustain T cell activation; functional OXPHOS is vital at this stage of differentiation and may explain why we see the lowest levels of m.3243A>G in cells which have undergone activation-induced mass clonal expansion. This is consistent with observations in m.3243A>G patient fibroblasts where restricting access to glucose and glutamine forces a reliance on pyruvate as a substrate, driving positive selection for wild-type and negative selection against variant mtDNA molecules(43). This selection is likely to be the result of selective depolarisation of mitochondria containing variant mtDNA, inhibiting the replication of variant mtDNA molecules and increasing wild-type mtDNA replication(43). In T cells, aged mitochondria are asymmetrically segregated into daughter cells with higher levels of autophagy, mitochondrial clearance and self-renewal upon T cell activation-related cell division(42). It is possible that a similar mechanism identifies mitochondria with higher mutation levels and that that both asymmetric segregation followed by selective mitophagy may contribute to the enhanced reduction in mutation levels that we observe in memory T cells. In our data, we see a higher mtDNA CN in cells of lymphoid origin in patients with higher m.3243A>G levels; this may be related to a higher mitochondrial mass, compensating for a biochemical defect.

The mTOR pathway has been shown to be chronically activated in m.3243A>G patient fibroblasts; inhibition of this pathway with rapamycin reduces fibroblast m.3243A>G levels(44). The mTOR pathway is also linked to activation-induced T cell mitochondrial biogenesis(45) and leucine, acting via this pathway plays a key role in T cell activation(46). This relationship is intriguing given that m.3243A>G disrupts mt-tRNA^Leu(UUR)^, and may imply that selection against m.3243A>G is linked to amino acid homeostasis. How this links to selection against m.8433A>G (within mt-tRNA^Lys^) is unclear. Therefore, further investigation into selection against other pathogenic mtDNA variants in mt-tRNA and protein coding genes, and the potential link to the mTOR pathway is merited.

Despite being well characterised in whole blood, genetic heterogeneity of mtDNA mutation level and the complex cellular composition of blood tissue have both previously hampered our understanding of the cellular mechanisms involved in the selection against mtDNA mutations. Here, we demonstrate the power of combining innovative single cell genomic technologies with high resolution cell phenotyping to gain further insight into the processes underlying blood cell mutation decline and the relationship between T cell development and mitochondrial function. Future studies into the strikingly high levels of negative selection in T cells, taking advantage of the rapid development of ‘omic technologies that can interrogate cellular pathways at the single cell level, will give us insights into the key processes involved in this mechanism. This has the potential to identify drug targets that could be useful in the treatment of mitochondrial disease, and to inform our understanding of the key role that mitochondria play in the differentiation of T cell lineages.

## Methods

### Participant recruitment

Twenty-two individuals (**Table S1**) carrying the m.3243A>G variant and four carrying m.8344A>G were recruited from the NHS Highly Specialised Service for Rare Mitochondrial Disorders in Newcastle upon Tyne, UK; seven healthy controls via the Dendritic Cell Homeostasis in Health and Disease study (08/H0906/72) and an additional 12 healthy control samples were obtained from the MAGMA study (17/NE/0015)(**Table S2**). All m.3243A>G patients underwent clinical assessment using the Newcastle Mitochondrial Disease Adult Scale (NMDAS)(47) to evaluate their disease phenotype and severity. None of these patients reported having any infective symptoms or taking any anti-microbial treatment in the preceding three weeks. This was confirmed by patients’ blood cell counts, which were all within normal ranges, apart from P1 who had a mildly reduced lymphocyte count (full blood count data presented in **Table S3**). Of the 22 m.3243A>G patients, we identified six, representing a range of ages (18-46 years) and whole blood .3243A>G level (9-60%), to take forward to single cell analysis.

### Sample Preparation

Whole blood samples (8-20ml, collected in EDTA) were separated via ficoll density gradient using Lymphoprep (Stem Cell Technologies) into three populations: peripheral blood mononuclear cells (PBMCs), platelets and granulocytes. Platelets were harvested from the plasma fraction and a granulocyte pellet was obtained by lysis of the red cell pellet using 20ml red cell lysis buffer (155mM ammonium Chloride, 12mM sodium bicarbonate & 0.1mM Ethylenediaminetetraacetic acid (EDTA)) Blood samples >15ml were prepared in two aliquots. granulocyte and platelet populations were pelleted and frozen at −80°C. PBMCs were washed twice with phosphate buffered saline (PBS), split into 3-8 1ml aliquots with freezing media (FBS + 10% DMSO) and stored as viable cells at −80°C.

### Cell Sorting and lysis

PBMC samples were stained with fluorescently labelled antibodies (**Table S4**) and sorted with a BD ARIA Fusion with a 70-micron nozzle according to gating strategy indicated (**Figure S1**).

### Bulk cell sorting

Up to 1000 cells of each cell type were sorted into Eppendorf tubes containing 25μl lysis buffer (500mM Tris-HCl, 1% Tween20, dH2O, 20mg/mL Proteinase K). mtDNA copy number (CN) calculations were adjusted accordingly for rare populations in which fewer cells were available (e.g CD34+ progenitor cells), Sorted samples were centrifuged to ensure all cells were submerged in buffer, lysed by incubating at 55°C for 2 hours and 10 minutes at 95°C and frozen at −20°C.

### Single cell sorting

Single cells were sorted into 15ul of lysis buffer (as above) in a 96-well plate. Each plate contained cells of a single sort gate and included a negative control well.

### Bulk m.3243A>G measurements

Patient m.3243A>G levels were determined via pyrosequencing using the Pyromark Q24 system as previously described and validated(48). Five control samples with known levels (15%, 46%, 70%, 0% and 0%) allowed for validation of each pyrosequencing run. If any of the control heteroplasmy readings differed from the known m.3243A>G level by more than +/-3%, the experiment was repeated. Primers used to target m.3243A>G in the initial DNA amplification reaction were obtained from IDT (Coralville, USA) (sequences according to GenBank Accession number NC_012920.1: 50biotinylated forward: m.3143-3163; reverse: m.3331-3353). And within the pyrosequencing reaction a reverse sequencing primer was used: m.3244-3258.

### mtDNA copy number

Mitochondrial DNA copy number was quantified in triplicate using real-time PCR, as previously described(6). Briefly, each 25μl population lysate was diluted 1:5 with nuclease free water and 5μl of each diluted sample was used per 15μl reaction. Primer and probe sequences relative to GenBank number NC_012920.1: forward: m.3485-3504; reverse: m.3553-3532; VIC-labelled MT-ND1 probe (m.3506-3529). Probes contained a non-fluorescent quencher and 3’ MGB moiety. Each 96-well plate contained a standard curve derived from six tenfold serial dilutions of a plasmid containing one copy of the ND1 target. Plasmid copies per μl for each dilution were determined using the DNA concentration of the plasmid sample and its molecular weight. R^2^ values were > 0.9992 and gradients fell between −3.264 and −3.449 for all standard curves. Mean threshold cycle (Ct), standard curves and the number of cells per sample (as counted by the cell sorter) were used to determine the absolute mtDNA copy number per cell for each sample. Within sample outliers and samples with Ct standard deviation > 0.3 or mean Ct > 30 were excluded.

A control DNA sample was included on each plate and used to standardise mtDNA copy number to control for inter-plate variation using the following formula: normalised mtDNA copy number = (absolute mtDNA copy number/on-plate control mtDNA copy number)*mean control mtDNA copy number.

### Single cell genomics

After cell lysis, 2μl of a unique barcoded forward primer (10μM; **Table S5**) was added to each well followed by the addition of 18μl containing high fidelity Platinum™ SuperFi II DNA Polymerase & buffer (Thermofisher), 10mM dNTP, 10μM reverse primer (5’ GGT TGG CCA TGG GTA TGT TG 3’), and nuclease free dH_2_O. Samples where then amplified via PCR according to the following cycling conditions: 30s at 98⁰C; 35 cycles of 7s at 98⁰C, 10s at 60⁰C, 30s at 72⁰C; 300s at 72⁰C.

PCR products from each plate were pooled (10μl per well) and unique Ion Xpress adaptors (numbers 46-80) were added, according to the Ion Plus Fragment library kit protocol (Life Technologies). 3μl product from each pooled plate was then combined to form the final library, which was quantified using an Aligent Technologies Bioanalyser. Ion Torrent Chip preparation was carried out using a final library concentration of 80pmol/l using an Ion Chef™, according to manufacturer’s instructions and the chip loaded onto an Ion S5™ sequencing machine within 15 minutes of reaction completion.

### Bioinformatics

Per plate BAM files were converted into fastq format using SAMtools v. 1.12(49). Reads containing well-specific barcode sequences in both the forward and reverse directions were extracted using Sabre(50); these were then realigned to a reference sequence (derived from RefSeq:NC_012920.1) using BWA v.0.7.17(51), producing SAM files, which were converted to well-specific pileup files using SAMtools v. 1.12(49). Variants were called using the mpileup2cns function in Varscan v2.3.9(52) with a minimum average quality of 28, a minimum variant allele frequency of 0.0001 and a minimum coverage of 1. Format fields from the vcf file were extracted using VCFtools v.0.1.16(53) and imported into R v.3.6.0(54) for downstream quality control and analysis.

Distributions of per-cell read depth per sequencing batch were inspected to determine an optimum lower threshold; cells with a read depth below this were excluded. To avoid the inclusion of wells containing multiple cells, we also excluded cells with a read depth greater than 1.5 times the upper boundary of the interquartile range. Three plates (P15: 8+ Naïve; P17: 34+ Progenitor; P18: 4+ CM) were excluded due to suspected contamination in the negative controls. 5732 cells passed quality control; median read depth was 1146 (IQR = 903, 2792, range = 201, 6975).

### Validation of technique

Amplicons which were not pooled for sequencing underwent a second PCR reaction using primers detailed in *Bulk heteroplasmy measurements.* This product was then sequenced using pyrosequencing and the m.3243A>G level measured. Comparison of the IonTorrent mutation level estimate and the pyrosequencing level estimate revealed the two techniques to be highly comparable (mean difference = 2.86%, median = 2.95%, with an overall difference in range of 0.07% to 6.87%) (**Figure S4**).

### Statistics

All statistical tests were performed in R v.3.6.0(54). Linear mixed models were performed using the nlme package(55) and included random intercepts for ID to account for variation due to individual. All graphs were produced using the ggplot2 package(56).

We considered p values < 0.05 to be significant and, where appropriate, p values were adjusted for multiple comparisons using the p.adjust function in R using the method=”fdr” option. Comparisons between two groups were performed using the wilcox.test function. Where shown, box-plots depict the median, 25th and 75th quantiles and whiskers extend from the hinge to the largest/smallest values no further than 1.5 * IQR from the hinge.

To determine the proportion of cells with near zero levels of m3243A>G, a Bayesian mixture model was fitted to the single cell mutation load data for each set of lineage observations for each subject. The model identified the spike in the data (near zero mutation load) and the proportion of cells which belonged to it. A mixture prior was placed on the proportion of cells belonging to the spike, imposing a probability of spike near zero mutation load.

The model is described as *Y∼πN01(μ,σ2)+(1−π)U(0,1), π∼π0δ0+(1−π0)U(0,1)*.

Where *N*01 denotes a normal distribution truncated on range [0,1] i.e. only values within this range can have a non-zero density and U0 denotes the Dirac delta function - a function whose value is zero everywhere except at zero, implying that π0 is the probability of no spike. The priors placed on the unknown parameters were: μ∼ U(0,0.2) σ∼Exp(5) π0∼U(0,1). Inference was carried out using the package rjags v.4.13(57).

### Study Approval

Written informed consent from participants was obtained prior to participation. All clinical investigations were evaluated according to the Declaration of Helsinki. Ethical approval was granted by the Newcastle and North Tyneside Research Ethics Committee (REC:19/LO/0117; “Understanding the decline in levels of the mitochondrial DNA mutation m.3243A>G within blood cell subtypes”). Additional control samples were obtained from the “Dendritic Cell Homeostasis in Health and Disease study” (REC:08/H0906/72) and the “MAGMA study” (REC:17/NE/0015).

## Data Availability

Underlying data and supporting analytic code for the manuscript can be accessed via https://github.com/sarahjpickett/SingleCellBloodM3243

## Author contributions

Designing research studies: MC, OR, SJP

Conducting Experiments and acquiring data: IGF, PM

Sample selection and collection: IB, GSG, YSN

Data analysis: IGF, JC, CL, SJP

Writing the manuscript: IGF, PM, MC, OR, SJP

All authors provided critical review of the manuscript.

## Acknowledgements

We would like to thank all individuals who agreed to participate in this study and the clinical, laboratory and research administration and support teams. We would also like to thank Helen Tuppen for help with optimising the IonTorrent single cell assay; Doug Turnbull and John Grady for helpful discussions leading up to this study; Rob Taylor and the Newcastle NHS Highly Specialised laboratory team for assistance with pyrosequencing assays; Andrew Schaefer, Catherine Feeney and Rhys Thomas for their role in recruiting and clinically assessing patients; Laura Brown, Clare Massarella for assistance with the ethical approval for this study; and Doug Jerry for clinical data management. This research made use of the Rocket High Performance Computing service at Newcastle University. We acknowledge the Newcastle University Flow Cytometry Core Facility (FCCF) for assistance with the generation of Flow Cytometry data. This work was supported by a Wellcome Career Re-entry Fellowship to SJP (204709/Z/16/Z); a MRC DiMeN DTP studentship to IGF, the Wellcome Centre for Mitochondrial Research (203105/Z/16/Z) and the UK NHS England Specialist Commissioners which funds the Highly Specialised Service for “Rare Mitochondrial Disorders of Adults and Children” in Newcastle upon Tyne. MC and PM are supported by Wellcome Trust Investigator Award 219562Z/19Z. Funding for open access charge: Charity open access fund (COAF). For the purpose of open access, the author has applied a CC BY public copyright licence to any Author Accepted Manuscript version arising from this submission. Figure 1, S3 & the graphical abstract were created with BioRender.com

