## Supplementary Information for "T cell differentiation drives the negative selection of pathogenic mtDNA variants"

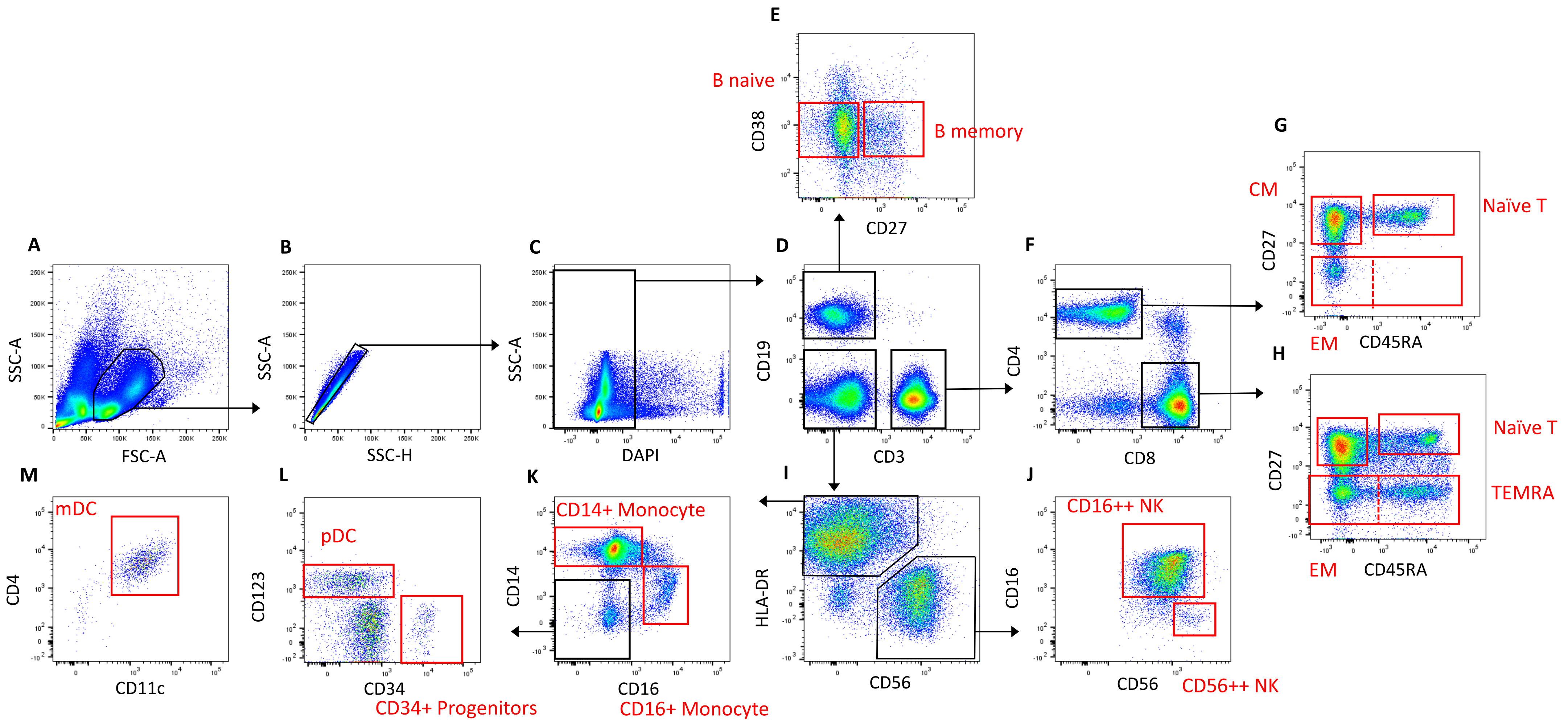
E
B naive
CD38
B memory
CD27
G
CM
Naïve T
CD27
EM
CD45RA
A
SSC-A
FSC-A
B
SSC-A
SSC-H
C
SSC-A
DAPI
D
CD19
CD3
F
CD4
CD8
H
Naïve T
CD27
TEMRA
EM
CD45RA
M
mDC
CD4
CD11c
L
pDC
CD123
CD34
CD34+ Progenitors
K
CD14+ Monocyte
CD14
CD16
CD16+ Monocyte
I
HLA-DR
CD56
J
CD16++ NK
CD16
CD56
CD56++ NK

**Figure S1:** ***Representative example of the FACS gating strategy for P15.*** *A) Cells gated on SSC-A and FSC-A. B) Doublets excluded with SSC-A and SSC-H. C) DAPI+ dead cells excluded. D) Cells split into CD19+ B Cells, CD3+ T Cells and CD19-3- cells. E) CD27-38medium Naïve Mature B cells (referred to from hereon as Naïve B cells) and CD27+38medium Memory B Cells (referred to from hereon as Memory B cells) gated to sort. F) T cells split by expression of CD4 and CD8. G&H) CD27+45RA+ Naïve T cells (referred to from hereon as Naïve T cells) and CD27- Memory T cells (referred to from hereon as Memory T cells) gated to sort from both CD4+ and CD8+ T cell fractions. In single cell investigations the memory T cell population was split into CD27-CD45RA- EM and CD27-CD45RA+ TEMRA and a separate CD27+CD45RA- CM population was sorted I) CD19-3- cells split into HLADR+56- Antigen Presenting Cells and HLADR-56+ Natural Killer Cells. J) NK cells split by expression of CD56 and CD16 for sort; CD56++CD16- NK Cells (referred to as CD56++ NK Cells) and CD56+CD16++ NK Cells (referred to as CD16++ NK Cells) K) CD14+ monocytes and CD16+ monocytes split for sort. L) CD123+ pDCs and CD34+ precursor cells split for sort. M) CD11c+ mDCs split for sort. Sort gates are bordered in red.*

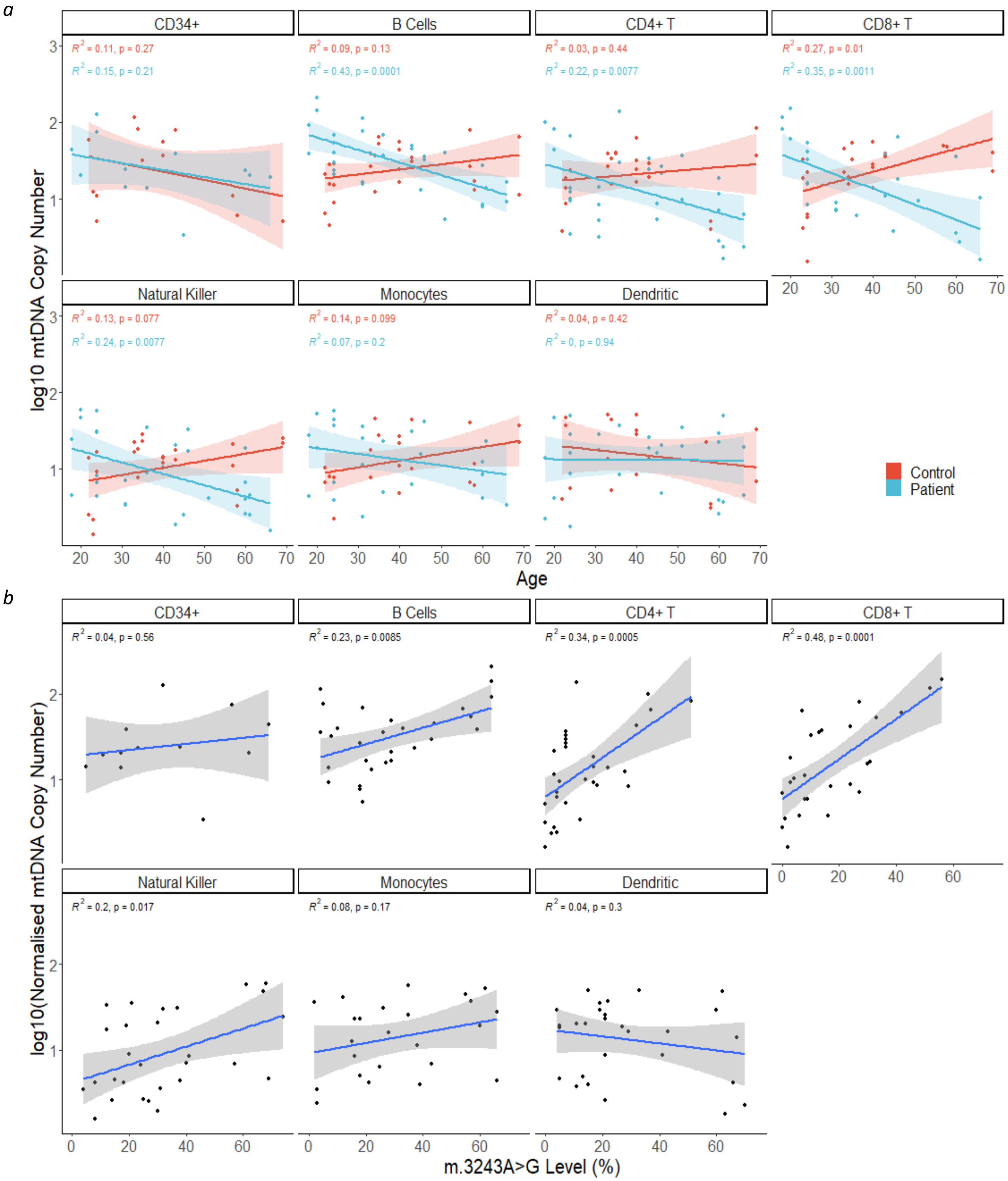
a
CD34+
B Cells
CD4+ T
CD8+ T
Natural Killer
Monocytes
Dendritic
R² = 0.11, p = 0.27
R² = 0.15, p = 0.21
R² = 0.09, p = 0.13
R² = 0.43, p = 0.0001
R² = 0.03, p = 0.44
R² = 0.22, p = 0.0077
R² = 0.27, p = 0.01
R² = 0.35, p = 0.0011
R² = 0.13, p = 0.077
R² = 0.24, p = 0.0077
R² = 0.14, p = 0.099
R² = 0.07, p = 0.2
R² = 0.04, p = 0.42
R² = 0, p = 0.94
log10 mtDNA Copy Number
Age
Control
Patient
b
CD34+
B Cells
CD4+ T
CD8+ T
Natural Killer
Monocytes
Dendritic
R² = 0.04, p = 0.56
R² = 0.23, p = 0.0085
R² = 0.34, p = 0.0005
R² = 0.48, p = 0.0001
R² = 0.2, p = 0.017
R² = 0.08, p = 0.17
R² = 0.04, p = 0.3
log10(Normalised mtDNA Copy Number)
m.3243A>G Level (%)

**Figure S2**: **mtDNA copy number in peripheral immune cell subsets.** *Linear regression lines, 95% CI, $R^2$ and p values are displayed on individual graphs. a) Relationship between age and log10 mtDNA copy number in cells from patients (blue; n=16) and controls (red; n= 14). Correlation coefficients and p values from linear models are shown on each panel; in controls, only CD8+ T cells show a significant increase with age ($\beta$=0.0149, SE=0.0053, p=0.0102). Patient cell subsets of lymphoid origin; B cells ($\beta$=-0.0163, SE=0.0036, p=0.0001), CD4+ T cells ($\beta$=-0.0151, SE=0.0053, p=0.0077), CD8+ T cells ($\beta$=-0.0202, SE=0.0055, p=0.0011) and NK cells ($\beta$=-0.0149, SE=0.0052, p=0.0077) have a negative relationship with age. To assess differences in the relationship between patients and controls, linear models with an interaction term between age and control/patient status were performed; there is a significant difference in slope between controls and patients for B cells (p=0.0001), CD4+ T cells (p=0.0216), CD8+ T cells (p=0.0001), NK cells (p=0.0018) and monocytes (p=0.0409). Interaction terms for other cells types were not significant (p>0.05). b) Comparison of relationship between m.3243A>G level and log10 mtDNA copy number in cell subsets in patients (n=16). In most cell types there is a trend for increasing mtDNA copy number as m.3243A>G levels increase; this relationship is significant for all cell types of lymphoid origin tested (B cells: $\beta$=0.0096, SE=0.0034, p=0.0085; CD4+ T cells: $\beta$=0.0229, SE=0.0059, p=0.0005; CD8+ T cells: $\beta$=0.0234, SE=0.0049, p=0.0001; NK cells: $\beta$=0.0106, SE=0.0041, p=0.0169).*

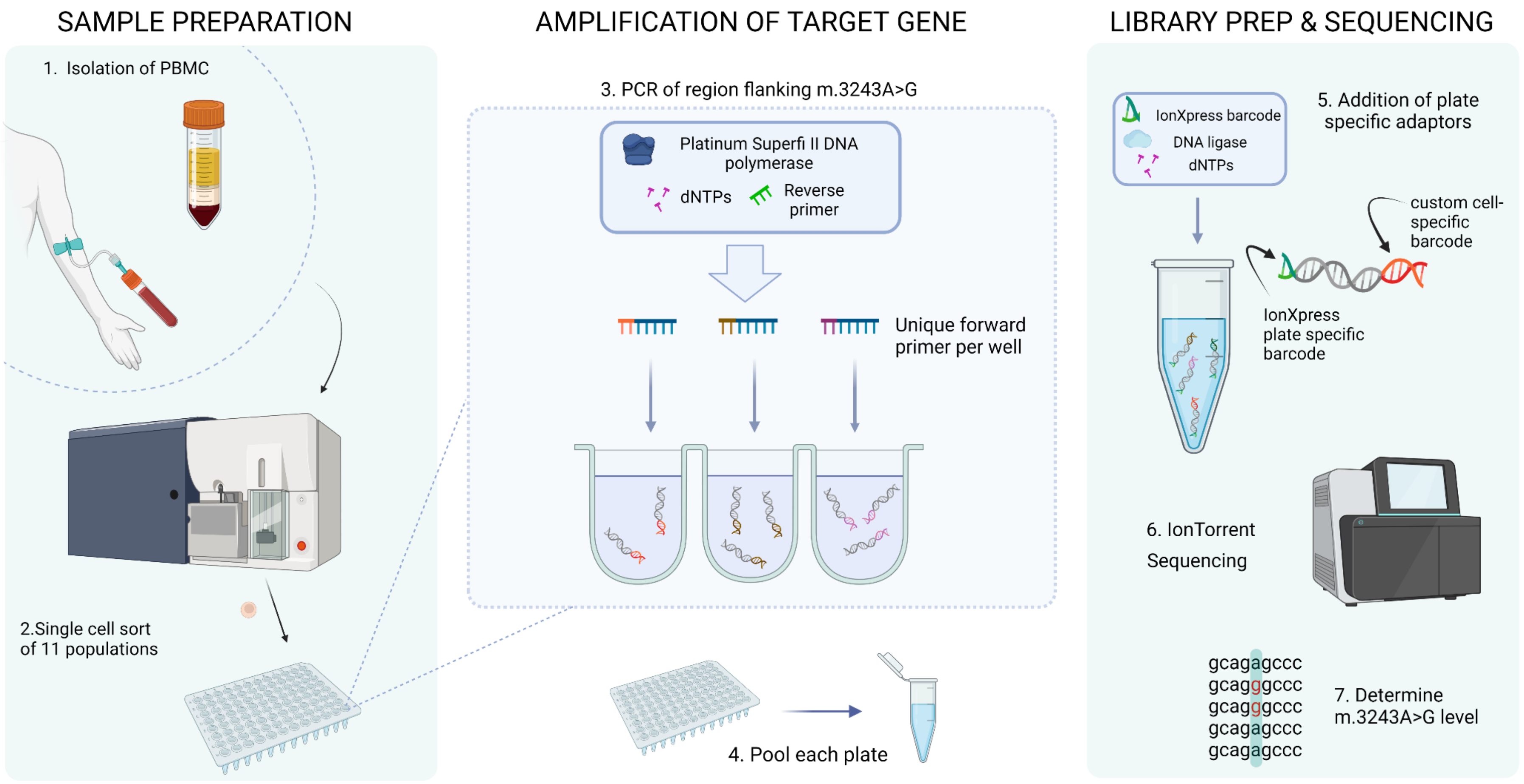

SAMPLE PREPARATION
1. Isolation of PBMC
2.Single cell sort
of 11 populations
AMPLIFICATION OF TARGET GENE
3. PCR of region flanking m.3243A>G
Platinum Superfi II DNA polymerase
dNTPs
Reverse primer
Unique forward primer per well
4. Pool each plate
LIBRARY PREP & SEQUENCING
IonXpress barcode
DNA ligase
dNTPs
5. Addition of plate specific adaptors
custom cell-specific barcode
IonXpress plate specific barcode
6. IonTorrent Sequencing
gcagagccc
gcagggccc
gcagggccc
gcagagccc
gcagagccc
7. Determine m.3243A>G level

**Figure S3: Determining m.3243A>G heteroplasmy in single cells using IonTorrent sequencing.** *1. PBMCs were isolated from whole blood via density centrifugation. 2. Single cells from 11 cell populations (CD34+ precursor cells, CD4+ naïve T cells, CD8+ naïve T cells, CD4+ CM, CD4+ EM, CD8+ CM, CD8+ EM, CD8+ TEMRA, naïve B cells, memory B cells and monocytes) were sorted into 96 well plates containing lysis buffer using FACS. 3. After cell lysis, the region surrounding mt.3243 was amplified from the whole cell lysate using well-specific barcoded forward primers and a common reverse primer. 4. Each plate of amplicons was then pooled. 5. Plate-specific IonXpress adaptors were added during library preparation. 6. Plates were then pooled to form the final library and sequenced on an IonTorrent S5 system. 7. After de-mulitplexing, single cell m.3243A>G level was determined using ratio of reads containing A:G at position m.3243. Created with BioRender.com*

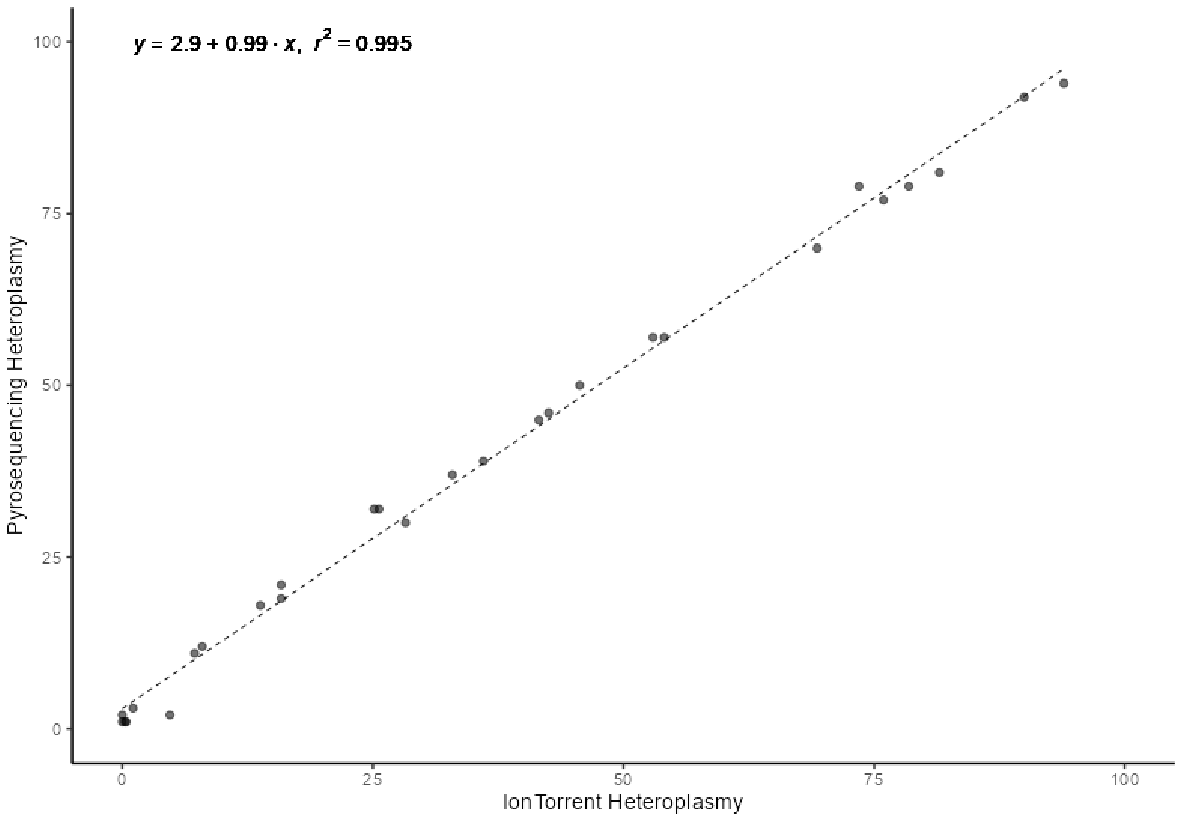


**Figure S4: Measurements of m.3243A>G levels were highly comparable between pyrosequencing and IonTorrent methods.**

| Study ID | Age | Sex | mtDNA variant | Clinical phenotype | Scaled total NMDAS | Most recent tissue level (%) | | | In single cell experiment |
|---|---|---|---|---|---|---|---|---|---|
| | | | | | | Blood (age) | Muscle | Urine | |
| P01 | 56-65 | M | m.3243A>G | Deafness, Diabetes mellitus, Cardiovascular involvement, Ptosis, Myopathy, Cerebellar ataxia, End-stage Renal Failure | 24.9 | 24 (57) | 80 | 84 | |
| P02 | 56-65 | F | m.3243A>G | Deafness, Diabetes mellitus | 16.6 | 21 (58) | N/A | 41 | |
| P03 | 46-55 | M | m.3243A>G | Diabetes mellitus, mild hearing deficit | 8.3 | 20 (38) | N/A | 67 | |
| P04 | 56-65 | M | m.3243A>G | Diabetes mellitus, Ptosis, Myopathy, Cerebellar ataxia, Dysphagia | N/A | 18 (59) | N/A | 73 | |
| P05 | 24-35 | M | m.3243A>G | Deafness | 4.1 | 6 (27) | 25 | 52 | |
| P06 | 46-55 | F | m.3243A>G | Deafness, chronic constipation | 9.3 | 13 (49) | N/A | 35 | |
| P07 | 56-65 | F | m.3243A>G | Impaired glucose tolerance, chronic constipation | 9.3 | 10 (55) | N/A | 57 | |
| P08 | 36-45 | M | m.3243A>G | Deafness, Diabetes mellitus, chronic kidney disease (focal segmental glomerulosclerosis) | 10.4 | 32 (40) | N/A | N/A | |
| P09 | 65+ | M | m.3243A>G | Deafness, Diabetes mellitus, Dysphonia/dysarthria, Cerebellar ataxia, Neuropathy | 32.2 | N/A | 67 | 66 | |
| P10 | 24-35 | M | m.3243A>G | Deafness, Myopathy, Chronic Constipation, Short stature | 17.6 | 59 (22) | N/A | 92 | y |
| P11 | 36-45 | F | m.3243A>G | Diabetes mellitus, Myopathy | 8.3 | 11(36) | 48 | 49 | |
| P12 | 24-35 | F | m.3243A>G | Deafness, Diabetes mellitus, Cardiovascular involvement, Short Stature, Renal transplant for focal segmental glomerulosclerosis | 18.6 | 36 (31) | N/A | 42 | |
| P13 | 24-35 | F | m.3243A>G | Mild hearing deficit | 7.3 | 31 (24) | N/A | 47 | |
| P14 | 56-65 | F | m.3243A>G | Deafness, CPEO, Cerebellar ataxia | 14.5 | 14 (60) | N/A | 74 | |
| P15 | 36-45 | M | m.3243A>G | Deafness, Diabetes Mellitus, Cerebellar Ataxia, Myopathy, Cardiovascular involvement | 45.2 | 23 (45) | N/A | 85 | y |
| P16 | 36-45 | F | m.3243A>G | Deafness | 4.1 | 23 (43) | 71 | 76 | |
| P17 | 46-55 | F | m.3243A>G | Myopathy, Deafness | 17.6 | 9 (46) | N/A | 64 | y |
| P18 | 24-35 | F | m.3243A>G | Deafness, chronic constipation | 14.5 | 36 (24) | N/A | 80 | y |
| P19 | 18-23 | F | m.3243A>G | Deafness, severe psychiatric involvement, migraines, cardiovascular involvement | 24.9 | 59 (20) | N/A | 70 | y |
| P20 | 25-35 | M | m.3243A>G | MELAS syndrome, dementia, chronic constipation | 51.8 | 42 (25) | N/A | N/A | |
| P21 | 65+ | M | m.3243A>G | Deafness, Myopathy, Ataxia, Neuropathy | N/A | 12(65) | N/A | 77 | |
| P22 | 18-23 | F | m.3243A>G | Deafness, Cardiovascular involvement, Myopathy, Short Stature | 24.5 | 60 (18) | N/A | 96 | y |
| P23 | 56-65 | F | m.8344A>G | Asymptomatic | 6.4 | 55 (58) | N/A | N/A | |
| P24 | 24-35 | M | m.8344A>G | MERRF syndrome, lipomatosis, cerebellar ataxia, deafness | 46.2 | 86 (24) | N/A | N/A | |
| P25 | 65+ | F | m.8344A>G | Myoclonus, Neuropathy, Hearing impairment, cerebellar ataxia | 10.7 | 65 (75) | N/A | 67 | |
| P26 | 24-35 | F | m.8344A>G | Ataxia, lipomatosis, mild hearing deficit | N/A | 67 (35) | N/A | N/A | |

**Table S1: Demographics and clinical details of the patient cohort**. CPEO: chronic progressive external ophthalmoplegia; MELAS: mitochondrial encephalomyopathy, lactic acidosis and stroke-like episodes; MERRF: myoclonic epilepsy with ragged red fibers; N/A: measurement not available.

| Study ID | Age | Sex |
|---|---|---|
| C01 | 65+ | M |
| C02 | 46-55 | M |
| C03 | 24-35 | M |
| C04 | 56-65 | M |
| C05 | 46-55 | M |
| C06 | 56-65 | M |
| C07 | 56-65 | M |
| C08 | 65+ | M |
| C09 | 18-23 | M |
| C10 | 18-23 | M |
| C11 | 18-23 | F |
| C12 | 24-35 | F |
| C13 | 24-35 | F |
| C14 | 36-45 | F |
| C15 | 36-45 | F |
| C16 | 24-35 | F |
| C17 | 36-45 | M |
| C18 | 24-35 | M |
| C19 | 24-35 | F |

**Table S2: Demographics of control cohort.**

| Patient | Gender | Age at Visit | Full Blood Count Data | | | | | | | | | | | | | |
|---|---|---|---|---|---|---|---|---|---|---|---|---|---|---|---|---|
| | | | WBC (x 10'9/L) | RBC (x 10'12/L) | RCDW (%) | Reticulocyte Count (x 10'9/L) | MCV (fL) | MCH (pg) | Haematocrit (L/L) | Haemoglobin (g/L) | Lymphocyte Count (x 10'9/L) | Basophil Count (x 10'9/L) | Eosinophil Count (x 10'9/L) | Monocyte Count (x 10'9/L) | Neutrophil Count ( x 10'9/L) | Platelet Count (x 10'9/L) |
| P1 | MALE | 56-65 | 10.13 | 3.97 | none | none | 96 | 30 | 0.381 | 119 | 0.34 | 0.03 | 0.03 | 0.52 | 9.21 | 248 |
| P2 | FEMALE | 56-65 | 6.66 | 4.87 | none | none | 88.9 | 29.4 | 0.433 | 143 | 1.95 | 0.06 | 0.27 | 0.61 | 3.77 | 349 |
| P3 | MALE | 46-55 | 8.12 | 4.69 | none | none | 97.4 | 33.5 | 0.457 | 157 | 3.1 | 0.07 | 0.28 | 0.64 | 4.03 | 275 |
| P4 | MALE | 56-65 | 10.15 | 4.56 | none | none | 94.7 | 32.5 | 0.432 | 148 | 2.84 | 0.13 | 0.68 | 1.19 | 5.31 | 226 |
| P5 | MALE | 24-35 | 6.4 | 5.61 | none | none | 86.6 | 30.5 | 0.486 | 171 | 1.52 | 0.06 | 0.21 | 0.67 | 3.94 | 297 |
| P6 | FEMALE | 46-55 | 7.96 | 4.74 | none | none | 88.4 | 28.3 | 0.419 | 134 | 1.41 | 0.05 | 0.21 | 0.92 | 5.37 | 369 |
| P7 | FEMALE | 56-65 | 7.05 | 4.77 | none | none | 92.7 | 30.8 | 0.442 | 147 | 2.05 | 0.08 | 0.25 | 0.72 | 3.95 | 335 |
| P8 | MALE | 36-45 | 6.88 | 4.02 | none | none | 86.8 | 30.3 | 0.349 | 122 | 2.34 | 0.02 | 0.22 | 0.63 | 3.67 | 204 |
| P9 | MALE | 65+ | 8.01 | 4.6 | none | none | 84.1 | 28.5 | 0.387 | 131 | 2.73 | 0.05 | 0.39 | 0.83 | 4.01 | 271 |
| P10 | MALE | 24-35 | 5.19 | 4.39 | none | none | 88.4 | 29.4 | 0.388 | 129 | 1.86 | 0.04 | 0.24 | 0.65 | 2.4 | 224 |
| P11 | FEMALE | 36-45 | 8.94 | 5.51 | none | none | 89.5 | 30.1 | 0.493 | 166 | 2.26 | 0.05 | 0.1 | 0.54 | 5.99 | 325 |
| P12 | FEMALE | 24-35 | none | none | none | none | none | none | none | none | none | none | none | none | none | none |
| P13 | FEMALE | 24-35 | 7.72 | 4.65 | none | none | 91.8 | 31.4 | 0.427 | 146 | 2.07 | 0.02 | 0.12 | 0.5 | 5.01 | 434 |
| P14 | FEMALE | 56-65 | 6.92 | 4.37 | none | none | 93.6 | 30.2 | 0.409 | 132 | 2.1 | 0.05 | 0.14 | 0.69 | 3.94 | 222 |
| P15 | MALE | 36-45 | 8.14 | 4.83 | none | none | 88 | 30.2 | 0.425 | 146 | 2.87 | 0.08 | 0.23 | 0.68 | 4.28 | 216 |
| P16 | FEMALE | 36-45 | 6.74 | 5.12 | none | none | 85.7 | 28.5 | 0.439 | 146 | 1.72 | 0.02 | 0.11 | 0.48 | 4.41 | 242 |
| P17 | FEMALE | 46-55 | 9.4 | 4.54 | none | none | 87.4 | 29.3 | 0.397 | 133 | 2.9 | 0.15 | 0.72 | 0.69 | 4.94 | 270 |
| P18 | FEMALE | 24-35 | 11.06 | 4.02 | none | none | 92 | 29.9 | 0.37 | 120 | 2.66 | 0.06 | 0.48 | 1.04 | 6.82 | 248 |
| P19 | FEMALE | 18-23 | 6.78 | 4.2 | none | none | 88.6 | 28.8 | 0.372 | 121 | 2.02 | 0.08 | 0.08 | 0.82 | 3.78 | 336 |
| P20 | MALE | 25-35 | 8.82 | 4.39 | none | none | 91.1 | 29.6 | 0.4 | 130 | 3.72 | 0.05 | 0.13 | 0.59 | 4.33 | 379 |
| P21 | MALE | 65+ | 7.14 | 4.27 | none | none | 93.4 | 30.9 | 0.399 | 132 | 1.29 | 0.05 | 0.35 | 0.77 | 4.68 | 223 |
| P22 | FEMALE | 18-23 | 8.14 | 4.7 | none | none | 85.1 | 28.1 | 0.4 | 132 | 2.04 | 0.07 | 0.2 | 0.75 | 5.08 | 278 |

**Table S3: Full blood count data**

| Bulk Sorting | | | | | |
|---|---|---|---|---|---|
| Antibody | Flourochrome | Clone | Company | Cat No | Panel |
| CD3 | FITC | SK7 | BD Biosciences | 345764 | 1&2 |
| CD4 | PE | SK3 | BD Biosciences | 345769 | 1&2 |
| CD8 | PerCP-Cy5.5 | SK1 | BioLegend | 344708 | 1&2 |
| CD11c | BUV395 | B-ly6 | BD Biosciences | 563787 | 1&2 |
| CD14 | BV650 | M5E2 | BD Biosciences | 563419 | 1&2 |
| CD16 | V500 | 3G8 | BD Biosciences | 561394 | 1&2 |
| CD19 | PE-CF594 | HIB19 | BD Biosciences | 562294 | 1&2 |
| CD27 | BV421 | O323 | BioLegend | 302823 | 1&2 |
| CD34 | PECy7 | 581 | BioLegend | 343516 | 2 |
| CD34 | BV605 | 581 | BioLegend | 343529 | 1 |
| CD38 | PECy7 | HB7 | BD Biosciences | 335825 | 1 |
| CD38 | BV605 | HB7 | BD Biosciences | 562665 | 2 |
| CD45 | A700 | HI30 | BioLegend | 304024 | 1&2 |
| CD45RA | APC-Cy7 | HI100 | BioLegend | 304127 | 1&2 |
| CD56 | APC | B159 | BD Biosciences | 555518 | 1&2 |
| CD123 | BV711 | 6H6 | BioLegend | 306030 | 1&2 |
| HLA-DR | BV786 | L243 | BioLegend | 307642 | 1&2 |
| DAPI | DAPI | n/a | Sigma Aldrich | D9542 | 1&2 |

**Table S4: Antibodies**

| Well | Barcode + forward |
|---|---|
| A01 | ACGATCGTGATAA GGC CTA CTT CAC AAA GCG |
| B01 | CTAGATCGTGTAA GGC CTA CTT CAC AAA GCG |
| C01 | GACTCGATCATAA GGC CTA CTT CAC AAA GCG |
| D01 | TGACTAGCTCTAA GGC CTA CTT CAC AAA GCG |
| E01 | ATGCTCAGCATAA GGC CTA CTT CAC AAA GCG |
| F01 | CGATCTGCATTAA GGC CTA CTT CAC AAA GCG |
| G01 | GATAGCTGACTAA GGC CTA CTT CAC AAA GCG |
| H01 | TCAGCTACGTTAA GGC CTA CTT CAC AAA GCG |
| A02 | AGTACGCATGTAA GGC CTA CTT CAC AAA GCG |
| B02 | CACGTCGATATAA GGC CTA CTT CAC AAA GCG |
| C02 | GTATCACGACTAA GGC CTA CTT CAC AAA GCG |
| D02 | TCGCAGTACTTAA GGC CTA CTT CAC AAA GCG |
| E02 | AGCGTCTGATTAA GGC CTA CTT CAC AAA GCG |
| F02 | CAGCATGTCTTAA GGC CTA CTT CAC AAA GCG |
| G02 | GTACTCATCGTAA GGC CTA CTT CAC AAA GCG |
| H02 | TCTGCAGCTATAA GGC CTA CTT CAC AAA GCG |
| A03 | ACTGTACTCGTAA GGC CTA CTT CAC AAA GCG |
| B03 | CGACAGCTATTAA GGC CTA CTT CAC AAA GCG |
| C03 | GTCATGCGTATAA GGC CTA CTT CAC AAA GCG |
| D03 | TAGTCGCATGTAA GGC CTA CTT CAC AAA GCG |
| E03 | ATCGATGACGTAA GGC CTA CTT CAC AAA GCG |
| F03 | CGATAGTCGTTAA GGC CTA CTT CAC AAA GCG |
| G03 | GAGCTGTATCTAA GGC CTA CTT CAC AAA GCG |
| H03 | TCTGATCGCATAA GGC CTA CTT CAC AAA GCG |
| A04 | AGCATCGTCTTAA GGC CTA CTT CAC AAA GCG |
| B04 | CTACGTCTAGTAA GGC CTA CTT CAC AAA GCG |
| C04 | GCTAGATGCTTAA GGC CTA CTT CAC AAA GCG |
| D04 | TCGAGTGCATTAA GGC CTA CTT CAC AAA GCG |
| E04 | ACGCTGACATTAA GGC CTA CTT CAC AAA GCG |
| F04 | CATACAGTGCTAA GGC CTA CTT CAC AAA GCG |
| G04 | GAGCACTAGTTAA GGC CTA CTT CAC AAA GCG |
| H04 | TGCATGTAGCTAA GGC CTA CTT CAC AAA GCG |
| A05 | AGTGATCGACTAA GGC CTA CTT CAC AAA GCG |
| B05 | CTGACATGCATAA GGC CTA CTT CAC AAA GCG |
| C05 | GTAGCAGATCTAA GGC CTA CTT CAC AAA GCG |
| D05 | TCACTATGCGTAA GGC CTA CTT CAC AAA GCG |
| E05 | ACTCGATACGTAA GGC CTA CTT CAC AAA GCG |
| F05 | CGCATGATCATAA GGC CTA CTT CAC AAA GCG |
| G05 | GCAGATCACTTAA GGC CTA CTT CAC AAA GCG |
| H05 | TCGACTAGTGTAA GGC CTA CTT CAC AAA GCG |
| A06 | ATCAGCGATGTAA GGC CTA CTT CAC AAA GCG |
| B06 | CTGTATGAGCTAA GGC CTA CTT CAC AAA GCG |
| C06 | GTGACTGTCATAA GGC CTA CTT CAC AAA GCG |
| D06 | TACGCTGCATTAA GGC CTA CTT CAC AAA GCG |
| E06 | AGCTGATGCATAA GGC CTA CTT CAC AAA GCG |
| F06 | CTATGCACTGTAA GGC CTA CTT CAC AAA GCG |
| G06 | GCTCATGTCATAA GGC CTA CTT CAC AAA GCG |

| | |
|---|---|
| H06 | TAGCGATCTGTAA GGC CTA CTT CAC AAA GCG |
| A07 | ACGTACTGCTTAA GGC CTA CTT CAC AAA GCG |
| B07 | CATAGCATCGTAA GGC CTA CTT CAC AAA GCG |
| C07 | CTGTGTGTGATAA GGC CTA CTT CAC AAA GCG |
| D07 | CAGTGAGAGCTAA GGC CTA CTT CAC AAA GCG |
| E07 | GTACATATGCTAA GGC CTA CTT CAC AAA GCG |
| F07 | GAGACTAGAGTAA GGC CTA CTT CAC AAA GCG |
| G07 | TACGCGTGTATAA GGC CTA CTT CAC AAA GCG |
| H07 | TGTCACTCATTAA GGC CTA CTT CAC AAA GCG |
| A08 | GCACATACACTAA GGC CTA CTT CAC AAA GCG |
| B08 | GCTCGTCGCGTAA GGC CTA CTT CAC AAA GCG |
| C08 | ACAGTGCGCTTAA GGC CTA CTT CAC AAA GCG |
| D08 | TCACACTCTATAA GGC CTA CTT CAC AAA GCG |
| E08 | TCACATATGTTAA GGC CTA CTT CAC AAA GCG |
| F08 | CGCTGCGAGATAA GGC CTA CTT CAC AAA GCG |
| G08 | ACACACAGACTAA GGC CTA CTT CAC AAA GCG |
| H08 | GCAGACTCTCTAA GGC CTA CTT CAC AAA GCG |
| A09 | TGCTCTCGTGTAA GGC CTA CTT CAC AAA GCG |
| B09 | GTGTGAGATATAA GGC CTA CTT CAC AAA GCG |
| C09 | CTCAGTGTGATAA GGC CTA CTT CAC AAA GCG |
| D09 | TGCGAGCGACTAA GGC CTA CTT CAC AAA GCG |
| E09 | GTCAGCTAGTTAA GGC CTA CTT CAC AAA GCG |
| F09 | AGATATCATCTAA GGC CTA CTT CAC AAA GCG |
| G09 | GTGCAGTGATTAA GGC CTA CTT CAC AAA GCG |
| H09 | TGACTCGCTCTAA GGC CTA CTT CAC AAA GCG |
| A10 | ATGCTGATGATAA GGC CTA CTT CAC AAA GCG |
| B10 | GACAGCATCTTAA GGC CTA CTT CAC AAA GCG |
| C10 | AGCGTCTGACTAA GGC CTA CTT CAC AAA GCG |
| D10 | TCGATATACGTAA GGC CTA CTT CAC AAA GCG |
| E10 | TCGTCATACGTAA GGC CTA CTT CAC AAA GCG |
| F10 | CGACTACGTATAA GGC CTA CTT CAC AAA GCG |
| G10 | GCGTAGACAGTAA GGC CTA CTT CAC AAA GCG |
| H10 | ACAGTATGATTAA GGC CTA CTT CAC AAA GCG |
| A11 | GTCTGATAGATAA GGC CTA CTT CAC AAA GCG |
| B11 | CTGCGCAGTATAA GGC CTA CTT CAC AAA GCG |
| C11 | TAGATCTCTGTAA GGC CTA CTT CAC AAA GCG |
| D11 | CTGATGCGCGTAA GGC CTA CTT CAC AAA GCG |
| E11 | CACTCGTGCATAA GGC CTA CTT CAC AAA GCG |
| F11 | TGACAGTATCTAA GGC CTA CTT CAC AAA GCG |
| G11 | GAGATACGCTTAA GGC CTA CTT CAC AAA GCG |
| H11 | ACGTGAGCTCTAA GGC CTA CTT CAC AAA GCG |
| A12 | ATAGAGAGTGTAA GGC CTA CTT CAC AAA GCG |
| B12 | CATAGAGAGATAA GGC CTA CTT CAC AAA GCG |
| C12 | ATCTCGAGATTAA GGC CTA CTT CAC AAA GCG |
| D12 | ACGATCACTCTAA GGC CTA CTT CAC AAA GCG |
| E12 | GATCGACTCGTAA GGC CTA CTT CAC AAA GCG |
| F12 | ATGCTCACTATAA GGC CTA CTT CAC AAA GCG |
| G12 | CGTGCACATCTAA GGC CTA CTT CAC AAA GCG |

| | |
|---|---|
| H12 | GACTGCACATTAA GGC CTA CTT CAC AAA GCG |

**Table S5: Barcoded custom forward primers for high throughput m.3243A>G genotyping.**

| Cell type | Patient (Age) | | | | | |
|---|---|---|---|---|---|---|
| | P22 (18-23 years) | P19 (18-23 years) | P10 (24-35 years) | P18 (24-35 years) | P15 (36-45 years) | P17 (46-55 years) |
| 34+Precursor | 0 | 0.001 | 0.001 | 0.216 | 0.358 | - |
| | (0.000, 0.000) | (0.000, 0.001) | (0.000, 0.008) | (0.134, 0.305) | (0.263, 0.461) | - |
| Monocyte | 0 | 0 | 0 | 0.194 | 0.462 | 0.741 |
| | (0.000, 0.000) | (0.000, 0.000) | (0.000, 0.022) | (0.115, 0.290) | (0.315, 0.640) | (0.64, 0.84) |
| 4+naive | 0.187 | 0.172 | 0.301 | 0.498 | 0.726 | 0.934 |
| | (0.105, 0.299) | (0.101, 0.261) | (0.215, 0.412) | (0.392, 0.594) | (0.615, 0.816) | (0.844, 0.987) |
| 4+CM | 0.404 | 0.47 | 0.734 | - | 0.882 | - |
| | (0.296, 0.5) | (0.394, 0.542) | (0.648, 0.824) | - | (0.737, 0.963) | - |
| 4+EM | 0.408 | 0.43 | 0.785 | 0.784 | 0.778 | 0.997 |
| | (0.308, 0.521) | (0.327, 0.521) | (0.704, 0.862) | (0.696, 0.866) | (0.668, 0.868) | (0.957, 1.000) |
| 4+TEMRA | - | - | - | 0.658 | - | - |
| | - | - | - | (0.549, 0.746) | - | - |
| 8+naive | 0.199 | NA | 0.439 | 0.406 | - | 0.774 |
| | (0.122, 0.299) | NA | (0.335, 0.554) | (0.304, 0.512) | - | (0.663, 0.874) |
| 8+CM | 0.334 | 0.508 | 0.813 | 0.601 | 0.86 | 0.996 |
| | (0.230, 0.431) | (0.411, 0.613) | (0.724, 0.882) | (0.497, 0.703) | (0.774, 0.941) | (0.942, 1.000) |
| 8+EM | 0.706 | 0.692 | 0.881 | 0.603 | 0.658 | 0.993 |
| | (0.598, 0.795) | (0.520, 0.866) | (0.800, 0.932) | (0.480, 0.698) | (0.538, 0.832) | (0.914, 1.000) |
| 8+TEMRA | 0.579 | 0.357 | 0.702 | 0.503 | 0.945 | 0.93 |
| | (0.481, 0.686) | (0.257, 0.497) | (0.603, 0.797) | (0.400, 0.617) | (0.839, 0.992) | (0.844, 0.990) |
| B Naïve | 0 | 0 | NA | 0.317 | 0.503 | 0.884 |
| | (0.000, 0.000) | (0.000, 0.000) | NA | (0.226, 0.399) | (0.410, 0.603) | (0.811, 0.939) |
| B Memory | 0.161 | 0 | NA | 0.375 | 0.663 | 0.891 |
| | (0.083, 0.255) | (0.000, 0.002) | NA | (0.273, 0.462) | (0.557, 0.748) | (0.764, 0.976) |

**Table S6: Peak estimates of the proportion of cells with a near zero m.3243A>G level.** *Dashes indicate missing data; 95% HDI are indicated in parentheses; NA indicates missing peak estimates due to bi/multimodal distributions*
